# Genetic loci associated with cocaine use identified using a novel approach to detect epistasis

**DOI:** 10.64898/2026.10.13.26365448

**Authors:** Michael R. Garvin, Andrew W. Bergen, Daniel M. Rosenblum, David Kainer, Sharlee Climer, James J Goedert, Peter J. Attia, Shihui Peng, Qiao Zhang, Stanley H. Weiss

**Affiliations:** Oregon Research Institute, Springfield, OR United States; Williwaw Biosciences, LLC, Clarkston, MI United States; Department of Medicine, Rutgers New Jersey Medical School, Newark, NJ United States; Rutgers Addiction Research Center, Rutgers Brain Health Institute, Rutgers University, New Brunswick, NJ United States; ARC Centre of Excellence for Plant Success in Nature and Agriculture, University of Queensland, Queensland, Australia; Department of Computer Science, University of Missouri St. Louis, St. Louis, MO United States; Anchor Mental Health, Catholic Charities of Washington, Washington, DC United States

**Keywords:** epistasis, copy number variation, addiction, cocaine, eQTL, HIV, CYP2D6, drug users

## Abstract

**Background:** Cocaine use disorder (CUD) is estimated to be 50% heritable, yet few genetic variants have been robustly associated with risk. During January 2021–June 2024, cocaine was involved in 30% of U.S. overdose deaths. Identifying genetic contributors to CUD could inform diagnostics and treatments. The “missing heritability” may stem from small sample sizes, models that inadequately account for small-effect common variants, and the exclusion of large-effect structural variants like copy number changes. Epistasis likely contributes but remains challenging to detect genome-wide.

**Results:** We focused on epistasis and haplotypes rather than individual SNPs by applying the custom correlation coefficient in *BlocBuster* to construct allele networks from eQTL SNPs. Testing these regulatory haplotype networks for association with cocaine use in 7,826 individuals with opioid use disorder identified 12 significant associations across two ancestries. For one haplotype, raw genotyping array intensity data revealed a copy number increase at the *CYP2D6* locus in individuals of European ancestry. In a subcohort of individuals who co-use cocaine and heroin (*i.e*., speedball), two haplotypes were also significantly associated with HIV infection. One of these involved GPR75, whose ligand, CCL5 (RANTES), is shared with CCR5, a coreceptor for HIV entry.

**Conclusions:** Our findings, integrated with long-read human pan-genomes, support a model in which non-functional copies of CYP2D6 disrupt dopaminergic, glutamatergic, and kynurenine pathways in the brain, contributing to cocaine use severity. The association of additional haplotypes with HIV status in speedball users highlights the genetic contribution to comorbid risk in high-exposure populations.

## Background

Cocaine is one of the most widely used psychostimulants, resulting in dependence in roughly 1% of the global population [1]. The associated public health burden has increased substantially, with 29,449 cocaine-involved overdose deaths reported in the United States in 2023. During January 2021–June 2024, cocaine was involved in 30% of reported overdose deaths, 79.1% of which also involved opioids [2,3]. As with many neuropsychiatric disorders, cocaine use disorder (CUD) is highly heritable (greater than 50%) [4–7], half of which has been attributed to common variation surveyed in genome wide association studies (GWAS)[1]. However, despite the substantial genetic contribution, conventional GWAS have had limited success in identifying replicated loci or biological pathways that account for the estimated heritability of CUD [8–11]. Identification of such pathways could allow for a personalized approach to diagnose, treat, and eventually prevent substance use disorder [12].

The “missing heritability” seen in CUD and other neuropsychiatric disorders is typically attributed to four sources: (1) small effects from many hundreds or thousands of common variants that are not captured by current models or sample sizes, (2) rare variants of large effect that are not presently included due to small sample sizes, (3) genomic structural variants such as copy number variation (CNV) of large effect that are not accurately identified with widely used short-read next generation sequencing, and (4) interaction effects (epistasis) that are difficult to model due to computational limitations when done at the level of the genome [13]. A key point is that the first two sources query a single source of heritability - effects of individual single nucleotide polymorphism (SNPs). Sources 3 and 4 are less commonly examined, but when they have a large effect size and are not rare, then they do not require large sample sizes to detect statistically significant associations, which would be advantageous for rare phenotypes or underrepresented populations.

One approach to analyze epistasis in the context of disease is the custom correlation coefficient (CCC) in the package BlocBuster [14], which generates pairwise correlations between all SNP alleles across the genome and then builds allele-based haplotype networks for subsequent testing. This highlights the fact that a SNP may be biologically relevant only in the context of a linked set (local, distant intra-chromosomal, or inter-chromosomal) and this signal would not be captured by single-site analyses such as GWAS. Indeed, this method has identified statistically significant haplotype networks that are associated with diseases and human traits that were missed by GWAS and can address phenotypic heterogeneity often seen with complex traits [14–16]. The main limitation is that the compute time of pairwise CCC increases exponentially with the number of SNPs and becomes difficult at scale [17]. One potential means to address this is to use expression quantitative trait loci (eQTL) SNPs from the genome as input rather than all SNPs. By using eQTLs we also ask a very specific question of genomic variation by effectively building gene expression networks (RNA) from SNPs (DNA). Additionally, the SNP-to-gene assignment is straightforward as it is based on the eQTL-associated expressed gene(s), and finally, because the association test with a phenotype is among a relatively small set of SNP-networks rather than each individual SNP, multiple testing correction is relaxed by orders of magnitude.

Here, we analyzed a unique cohort recruited from medication-assisted treatment centers for opioid dependence for genomic associations with cocaine use, which was reported only among some participants. Conventional single-variant GWAS produced limited and phenotype-dependent results: analysis of cocaine-use frequency identified no genome-wide-significant variants, whereas analysis of cocaine use as a binary phenotype identified two genome-wide-significant variants in DPYD within the African-ancestry subgroup. In contrast, our eQTL-restricted allele-network approach identified twelve multilocus haplotypes significantly associated with cocaine use, including three in those of European ancestry and nine in individuals of African ancestry. The strongest European-ancestry association involved a four-SNP haplotype in the *CYP2D6* region. Homozygosity for the *AACG* haplotype was associated with increased risk, whereas the *AACG/GGTA* diplotype was depleted among affected individuals, indicating a non-additive diplotype effect. The *AACG* homozygous association was subsequently validated in an independent cocaine-dependence cohort. Genotyping-array intensity data suggested that the risk haplotype was also associated with copy-number variation at *CYP2D6*. Analysis of recently reported haplotype-resolved long-read genomes indicated that the risk haplotype commonly occurs on structurally complex chromosomes containing nonfunctional *CYP2D6* copies, suggesting that increased DNA copy-number signal may correspond to reduced rather than increased CYP2D6 function. Further analyses of linked structural variation at the locus implicate the dopaminergic, glutamatergic, and kynurenine pathways.

## Results

### GWAS of Cocaine Use

The GRAF-POP approach [18] identified four population subgroups with sample sizes large enough for analysis (Table S1, Methods). GWAS for the frequency of cocaine use phenotype within each subpopulation failed to identify any significant loci after Bonferroni correction for the 714,238 tests (7 × 10^-8^, Figures S1-S4, Table S2a). The analysis identified suggestive risk loci (*p* < 1×10^-5^) in the European ancestry subpopulation (1CEU = 12), and in the two Hispanic ancestry subpopulations (5HS1 = 8, 6HS2 =9). No risk loci were identified in the African American (4AFA) subpopulation. In the African American subgroup, GWAS of cocaine use as a binary phenotype identified two genome-wide-significant variants, both mapping to *DPYD* (*P* < 3 × 10^^−8^; Table S2b). *DPYD* has previously been associated with several substance-use phenotypes [19,20]. The gene encodes dihydropyrimidine dehydrogenase, the rate-limiting enzyme in fluoropyrimidine metabolism, and clinically relevant variation in *DPYD* influences fluoropyrimidine toxicity and dosing. Importantly, the frequencies and functional consequences of *DPYD* variants differ among ancestry groups, emphasizing the need for ancestry-inclusive pharmacogenomic studies [21].

### eQTL-SNP networks associated with cocaine use

Of the 714,238 SNP loci on the array, 49,393 were known eQTLs according to GTEx or ORegAnno. Pairwise CCC of the eQTL SNPs were performed on each of the four subpopulations (1CEU, 4AFA, 5HS1, and 6HS2), producing 6719, 1828, 7, and 27 allele networks respectively for subsequent testing. The 5HS1 and 6HS2 produced few networks due to the need to increase the custom correlation coefficient threshold given that small sample sizes can increase the chances of spurious correlations (see Methods). A total of twelve eQTL CCC-allele networks were significantly associated with cocaine use after correction for multiple tests (Fig.1, Table S3); three in subpopulation 1CEU and nine in subpopulation 4AFA.

**Fig. 1.**
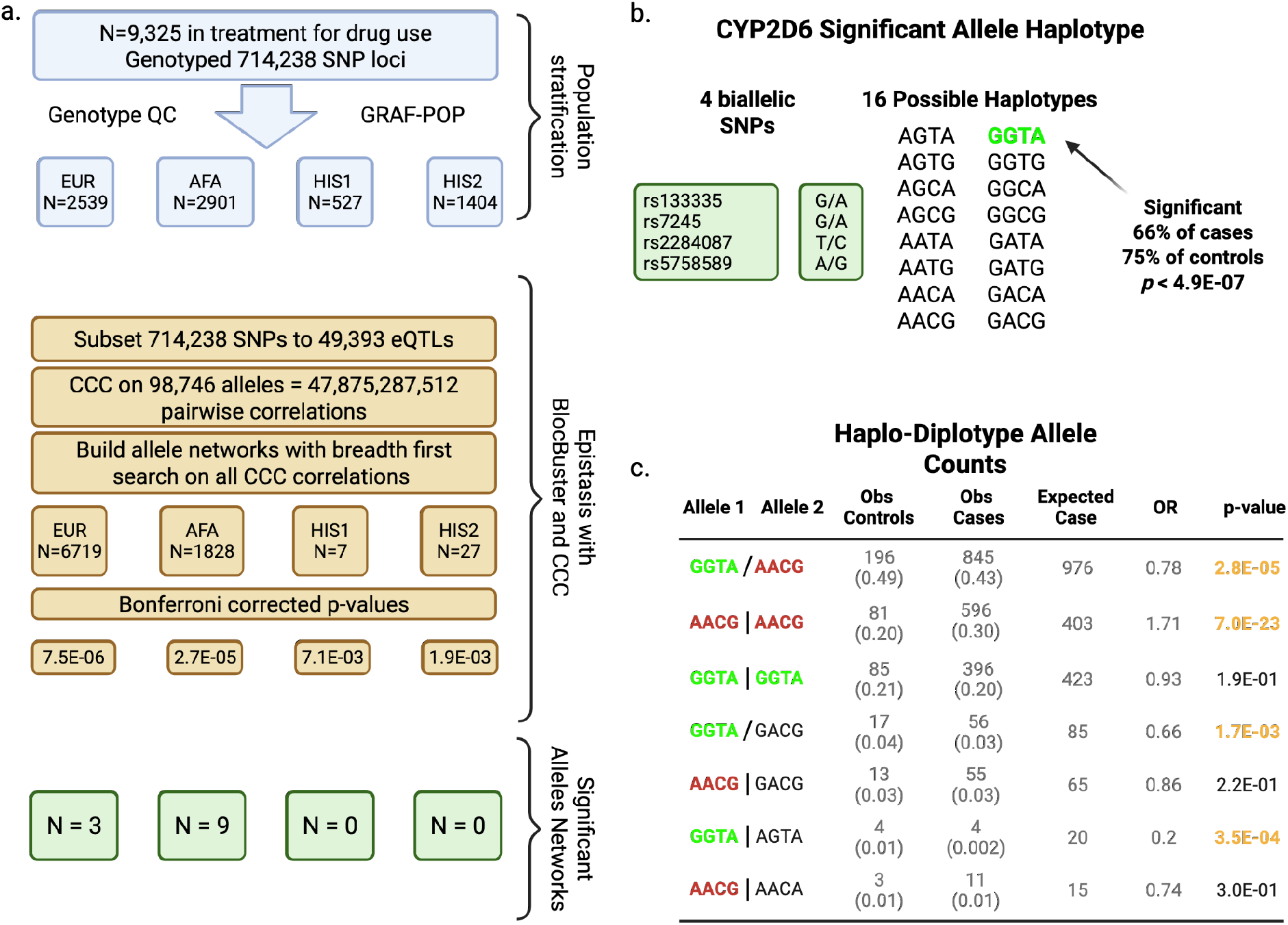
Summary of BlocBuster analysis. (a) After accounting for ancestry and genotype quality (light blue), networks were built for each GRAF-POP-defined subpopulation and then tested for differences between case and control (orange boxes). The number of networks tested for each subpopulation is provided. Note the Bonferroni corrected p-value thresholds are at least two orders of magnitude lower than a standard GWAS significant p-value threshold of 5 × 10 ^-8^. Three significant networks were found in the European subpopulation and nine in the African American subpopulation. (b) The four biallelic SNPs in one of the significant CCC subnetworks can produce up to sixteen different haplotypes. Significant haplotype shown in green is potentially protective as it is found at higher frequency in controls. Notably, an effect size of this magnitude could be captured at 5.5 × 10^-8^ with a GWAS having a sample size of 2,800 individuals. (c) Phased haplotypes for the significant locus in (b), with association tests stratified by diplotype. AACG homozygotes are significantly enriched among cases, while AACG/GGTA heterozygotes are significantly depleted, and GGTA homozygotes show no deviation from expectation. This pattern is consistent with an overdominant, non-additive model in which the reference GGTA haplotype buffers the pathogenic effect of AACG. EUR = European ancestry; His = Hispanic ancestry, AFA = African American ancestry. A “|” indicates phasing is known and “/” phase was predicted. Numbers in parentheses are frequencies. OR = Odds Ratio.

Significance thresholds for multiple tests differed based on the number of allele networks for each ancestry group and corresponded to 7.4 × 10^-6^, 2.7 × 10^-5^, 7.1 × 10^-3^, and 1.9 × 10^-3^ for 1CEU, 4AFA, 5HS1, and 6HS2 respectively.

Two of the three significant allele networks for subpopulation 1CEU regulate genes with known links to cocaine-related biological pathways and the third is indirectly implicated (Table 1, Table S3a). The G-protein coupled receptor GPR162 has been shown to bind the ligand, <u>c</u>ocaine- and <u>a</u>mphetamine-released transcript (CART) [22], which as the name suggests, was found to be upregulated in response to cocaine [23]. The protein encoded by *CYP2D6* appears to be the target of the second cocaine-related gene based on an analysis of GTEx associations. Although the most widely known role of CYP2D6 is metabolism of xenobiotic compounds in the liver, it can produce dopamine (cocaine binds to the dopamine transporter and blocks the reuptake of dopamine into the presynaptic neuron) through an alternative pathway [24]. The third allele network affects the expression of the glutamate transporter SLC25A22. Although this neurotransmitter is not directly involved with the immediate effects of cocaine administration, glutamate has been linked to later stages of addiction [25,26].

**Table 1.** Summary of significant CCC allele haplotypes with direct links to cocaine and dopamine pathways.

| Locus | Ancestry | eQTL prediction | Dopamine-related | HIV-linked | Architecture |
| --- | --- | --- | --- | --- | --- |
| CYP2D6 | EUR | ▲ | ● | – | Overdominance |
| GPR162/CD4 | EUR | ▼ | ● | ● | Overdominance |
| UNC13C | AFA | ▲ | ● | – | Intra-locus epistasis |
| GPR75 | AFA | ▲ | – | ● | Allele dominance |
| EPHB1 | AFA | ▼ | ● | – | Haplotype-specific dominance |
| SLC44A4/EHMT2 | AFA | ▼ | – | ● | Recessive-like |
| SERPINE1/ACHE | AFA | ▼ | ● | ● | Context-dependent dominance |

For subpopulation 4AFA, there were two networks known to be related to cocaine signaling pathways (Table 1, Table S3a). The first is a two-SNP allele haplotype that is associated with increased expression of the gene *UNC13C*, also known as *Munc13*, which modulates and coordinates synaptic vessel neurotransmitter release including dopamine [27,28] and is differentially expressed in response to cocaine in animal models [29].

The second network alters the expression of the tyrosine kinase receptor EPHB1, which was found to be upregulated in the brains of mice that were exposed to cocaine and the synthetic neurotoxin oxidopamine [30]. Three other allele haplotype networks affected the expression of genes that were indirectly linked to addiction and cocaine use. Two (SLC44A4 and ACHE) function to regulate acetylcholine. As with glutamate, this neurotransmitter has been implicated in longer term addiction processes [31].

### Dosage effects of phased diplotypes and epistasis on cocaine, HIV, and speedballing

In an additive genetic model, each risk allele contributes independently to disease risk. In contrast, epistasis arises when genetic variants interact such that the phenotypic effect of an allele depends on its genetic context. These interactions manifest as non-additive effects at the genotype or haplotype level. In this study we first identified statistically significant alleles based on presence–absence patterns. We then extended this framework by explicitly constructing phased haplo-diplotypes defined by the CCC-identified risk allele (see Methods). Diplotypes were formed by pairing the two haplotypes carried by each individual. This approach enables direct evaluation of dosage and pairing effects among CCC-identified regulatory alleles, rather than relying solely on conventional population-based phased haplotypes. Using this framework, we observed widespread non-additive genetic effects across multiple loci that deviated markedly from additive or dosage-based expectations for cocaine use (Table S4a, S4b).

Our initial focus was the non-additive effects on cocaine use, however, several of the genes linked to the regulatory CCC-allele haplotypes were also linked to HIV. For example, one of the CCC-alleles regulates *GPR162* expression, as well as its neighboring gene, the HIV receptor CD4. Similarly, GPR75 is the receptor for CCL5, which also binds the known HIV co-receptor CCR5 [32,33]. Finally, the regulatory CCC network for the choline transporter SLC44A4 also affects the expression of *EHMT2*, which has been linked to HIV latency [31,34]. Given these links, here we tested each of the diplotypes first for the association of cocaine use and then for HIV presence among all individuals, and the joint phenotype of HIV infection and speedballing, which is known to increase the risk of HIV infection in some populations (Table S4c, S4d).

At *CYP2D6*, haplo-diplotype analyses revealed a strong non-additive effect consistent with overdominance for cocaine use in the European ancestry subpopulation. Specific heterozygous haplotype combinations were depleted among cases (e.g. *AACG/GGTA*), while one homozygous haplotype (*AACG/AACG*) was significantly enriched, indicating clear deviation from additive genetic models. Notably, we confirmed the significant association of the *AACG/AACG* haplo-diplotype in a subset of individuals from the Yale-Penn cohort of cocaine users available in the study “Substance Dependence GWAS in European- and African - Americans” from the NIH database of genotypes and phenotypes (dbGAP, Table S4e). We performed the same analysis in a third cohort of individuals in the “Gene-Environment Interactions in COCCaINE Use Disorder: Collaborative Case-Control Initiative in Cocaine Addiction” also available on dbGAP. Only three of the four SNPs for the haplotype were present on the array in that latter study, and in this case, the homozygous diplotype (*AAC/AAC*) was not validated. However, when the analysis was reduced to only those who smoked, which more closely resembled the Weiss cohort, effect size was similar to that of Yale-Penn and statistically significant before Bonferroni correction (*p* < 0.05).

In order to test the robustness of the significant CCC allele (*GGTA*) at *CYP2D6* in the Weiss cohort, we divided the cases and controls into 70/30 splits and repeated the CCC BlocBuster analysis 100 times. The *GGTA*-containing *CYP2D6* bloc was recovered in all 100 random partitions (100/100), and the direction of effect was consistent across every analysis, with *GGTA* showing a protective association in all 100 partitions. The estimated relative risk was also highly stable across resamples (mean RR = 0.874, median RR = 0.875; range = 0.830–0.925). The association reached nominal statistical significance (*p* < 0.05) in 97 of 100 partitions and *p* < 0.01 in 81 of 100 partitions. In the three partitions that did not reach *p* < 0.05, the direction of effect remained unchanged (RR = 0.913–0.925), indicating reduced statistical strength rather than reversal of the association. The median *p*-value across the 100 analyses was approximately 0.0027.

Also in the European ancestry subpopulation, haplo-diplotypes at the *GPR162* locus demonstrated non-additive behavior consistent with haplotype-dependent overdominance, characterized by asymmetry between homozygous haplotypes and selective enrichment or depletion of specific heterozygous combinations. Only three of the four SNPs were available for the Yale-Penn cohort and comparison to the same 3-SNP haplotypes in Weiss revealed that in both, the *ACG/ACG* haplo-diplotype was significantly enriched in cases (Table S4e).

Haplo-diplotype analysis of the final locus in the European ancestry subpopulation at *SLC25A22* revealed extensive haplotype-dependent non-additive effects, with both enrichment and depletion of specific heterozygous and homozygous configurations. Unlike *CYP2D6*, no single heterozygous diplotype showed a consistent advantage. Instead, cocaine use depended on the specific pairing of long haplotypes, consistent with intra-locus epistasis across a multi-variant regulatory region. For the Yale-Penn cohort, the sample sizes were too small in those of European ancestry given the large number of haplotypes and therefore were not analyzed.

The nine loci that were significant in the Weiss cohort also demonstrated significance in the African ancestry subpopulation in the Yale-Penn cohort. At *GPR75*, haplo-diplotype analysis identified strong non-additive effects characterized by heterozygote depletion and asymmetric homozygote risk. The *CC/CC* configuration was significantly enriched among cases of cocaine use, whereas *TT/TT* and most heterozygous diplotypes were depleted, consistent with allele-dependent dominance rather than additive effects.

At *EPHB1*, haplo-diplotype analysis revealed allele-dependent non-additive effects with strong diplotype specificity. While the *TTGG/TTGG* homozygous configuration was significantly depleted among cases, multiple heterozygous haplotype combinations were either enriched or depleted relative to expectation. This bidirectional pattern is consistent with haplotype-specific dominance rather than uniform heterozygote or dosage effects.

At the *SLC44A4* locus, use of cocaine was dominated by a single risk configuration. The GC/GC homozygote was markedly enriched among cases, whereas *CT/CT* and *GC/CT* diplotypes were depleted, consistent with a homozygote-driven (recessive-like) architecture.

Finally, at *ACHE*, haplo-diplotype analysis revealed strong context-dependent non-additive effects. While the *GCT/GCT* configuration was significantly depleted and *TTC/TTC* showed modest enrichment, a single heterozygous configuration (*TTC/TCT*) was markedly over-represented among individuals that used cocaine. This pattern is consistent with haplotype-specific dominance with context-dependent heterozygote effects.

None of the haplo-diplotypes showed significant associations with HIV infection alone, but two demonstrated significant association for the joint phenotype of HIV infection and speedballing. At the *GPR75* locus, homozygous diplotypes showed no deviation from expectation, but a single heterozygous configuration (*CC/TC*) was significantly enriched among speedballing cases. Likewise, the S*LC44A4/EHMT2* locus effect was context-dependent. While the most common homozygous diplotypes showed no deviation from expectation, a single heterozygous configuration (*GC/GT*) was significantly enriched among those who speedball. These patterns are consistent with conditional epistasis, in which specific allele combinations influence risk only under combined substance exposure. Interestingly, in the seven loci that were able to be tested in the Yale-Penn and COCCaINE cohorts in those of African ancestry, tests were significant, but all were either the same haplo-diplotypes but in the opposite direction between cases and controls, or different haplo-diplotypes (Table S4e).

### GWAS Catalog associations with CCC-SNPs and Genes

To contextualize our findings with respect to existing single-variant GWAS literature, we queried the NHGRI–EBI GWAS Catalog for SNPs overlapping SNP loci highlighted in our analyses (Table S3b, S3c). While no cocaine- or psychostimulant-specific traits were reported, several variants mapped to phenotypes that are biologically and clinically relevant to addiction-related processes, including anorexia nervosa, major depressive disorder, attention-deficit/hyperactivity disorder, insomnia, and smoking initiation. In addition, many SNPs were associated with anthropometric and metabolic traits (*e.g*., body mass index, body fat distribution, glucose-related measures), as well as pharmacologic response and drug-metabolism phenotypes, consistent with the known involvement of metabolic, dopaminergic, and serotonergic pathways in substance use disorders.

These associations derive from marginal single-SNP effects identified in large population-based cohorts and therefore are not expected to directly recapitulate the haplotypic and interaction-based signals identified by the CCC/BlocBuster framework. Nonetheless, the convergence of psychiatric, addiction-related, metabolic, and pharmacologic phenotypes across these loci highlights their pleiotropic nature and is consistent with a model in which these genes participate in broader regulatory networks influencing reward processing, energy balance, and neuropsychiatric vulnerability.

### Potential causative associations with CYP2D6 AACG CCC-risk allele

Because the CCC analysis was restricted to eQTL-associated SNPs, the primary inferred mechanism underlying the observed associations is altered gene regulation. However, the *CYP2D6* locus is characterized by extensive structural complexity, including well-documented copy number variation (CNV), which provides an additional mechanism by which mRNA abundance may vary because the gain or loss of a gene copy can change the mRNA levels rather than simply due to increased or decreased transcription. We therefore examined genomic variation at this locus in greater detail.

The *AACG* haplotype is associated with increased *CYP2D6* expression relative to the reference *GGTA* haplotype across multiple tissues in GTEx (Fig. 2; Table S5). As expected given that CCC haplotypes were constructed from eQTLs, the direction of effect was consistent across all four constituent SNPs. For each SNP defining the *AACG* haplotype (rs133335 A, rs7245 A, rs2284087 C, rs5758589 G), the *AACG* allele was associated with higher *CYP2D6* expression relative to the alternate allele.

**Fig. 2.**
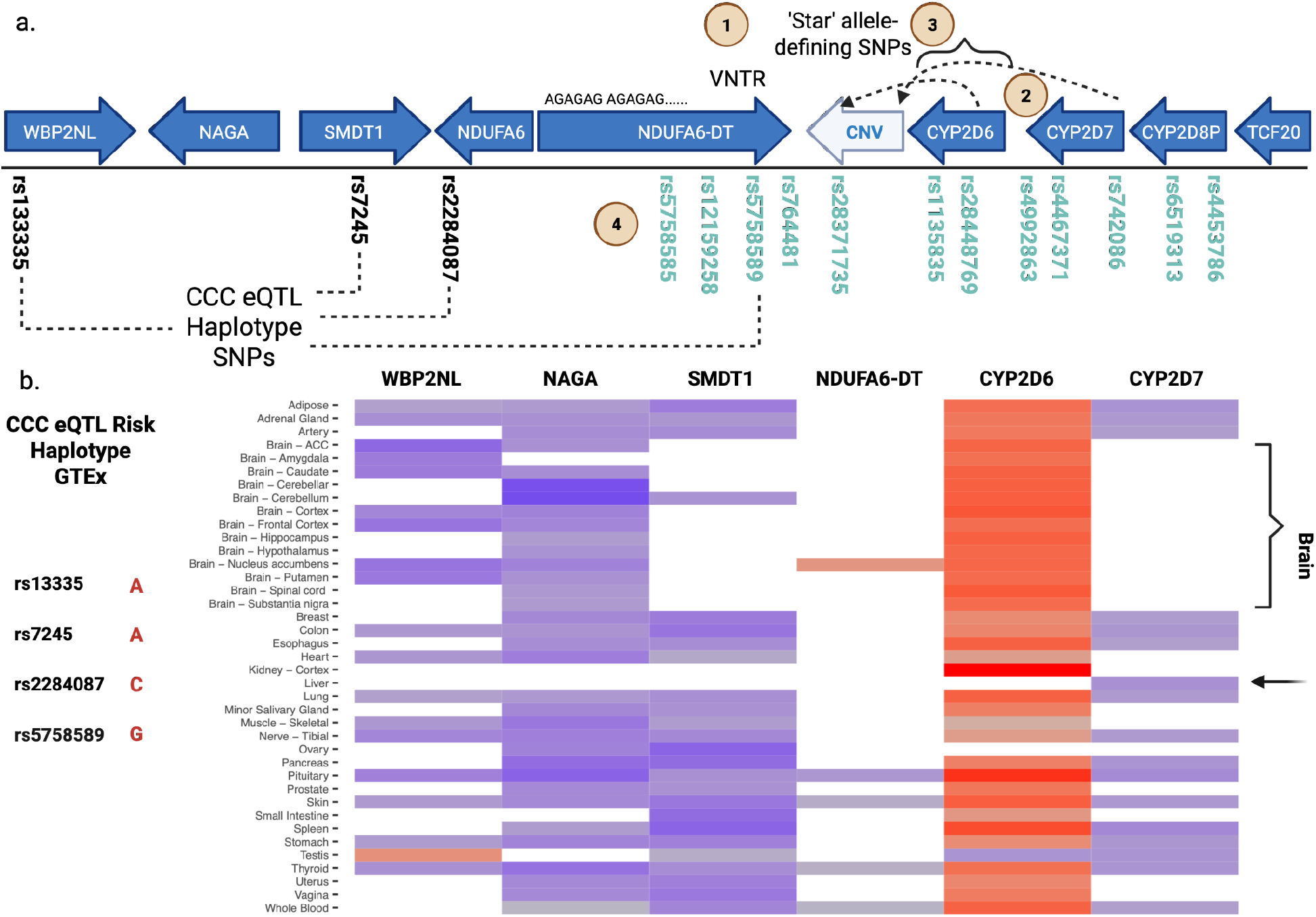
(a) Gene model of the CYP2D6 locus analyzed in this study. The four highlighted SNPs define haplotypes significantly associated with cocaine use in the CCC analysis. Additional genomic features evaluated for linkage with CCC-derived haplotypes are indicated: (1) a VNTR within NDUFA6-DT overlapping annotated transcription factor binding sites, (2) copy number variation affecting CYP2D6/CYP2D7, (3) SNPs defining star alleles or associated with altered CYP2D6 activity, and (4) SNP array markers used for raw intensity (Log R Ratio) analysis (see Fig. 4). (b) Normalized effect sizes (NES) for GTEx eQTLs corresponding to SNPs defining the AACG haplotype relative to the alternate GGTA haplotype. Red indicates increased expression associated with the AACG allele, blue indicates decreased expression, and gray indicates no significant effect. Notably, no eQTL effects were detected in liver despite the high baseline hepatic expression of CYP2D6. In contrast, regulatory effects were observed across multiple non-hepatic tissues, including several brain regions.

Notably, these eQTLs showed no detectable effect on *CYP2D6* expression in liver, despite this tissue exhibiting the highest baseline expression of *CYP2D6*. In contrast, the *AACG* haplotype was consistently associated with increased expression across multiple brain regions, suggesting that this locus primarily influences central nervous system regulation rather than hepatic expression.

In addition to its eQTL effects, the *AACG* haplotype overlaps variants annotated as splice quantitative trait loci (sQTLs). This observation is concordant with the finding that the *AACG* haplotype is strongly linked to rs3892097, the defining variant of the *CYP2D6*4* haplotype. Rs3892097 disrupts a splice acceptor site, resulting in a frameshifted transcript and loss of functional protein. The co-occurrence of eQTL and sQTL annotations at this haplotype therefore indicates coordinated regulatory effects on both transcript abundance and transcript integrity.

One explanation for the association between the four-SNP haplotype (rs133335, rs7245, rs2284087, rs5758589) and altered *CYP2D6* regulation is linkage disequilibrium with causative genomic features within or near the *CYP2D6* locus. This region contains extensive segmental duplications and complex structural variation that cannot be reliably resolved using SNP arrays or short-read sequencing. Direct detection of CNV and structural variants was therefore not possible in our cohort.

To address this limitation, we leveraged long-read, haplotype-resolved chromosomal assemblies. Using 148 phased assemblies from two independent population studies [31,32], we annotated structural and sequence features segregating in the general population and identified multiple variants strongly associated with the *AACG* haplotype (Fig. 2c; Tables S6–S9).

1. A variable number tandem repeat (VNTR) within *NDUFA6-DT* consists of 44 tandem 6-bp repeats overlapping transcription factor binding sites for STAT3, CEBPB, SMC1, and MAF [33]. These factors bind in response to interferon gamma (IFNG) and function as repressors [34,35]. Deletions overlapping these binding regions were present in all European-ancestry haplotypes carrying *AACG*, whereas *GGTA* haplotypes retained the intact sequence. Loss of repressive elements is consistent with increased *CYP2D6* expression observed for the *AACG* allelic class.
2. CNV at the *CYP2D6/CYP2D*7 locus revealed duplications and deletions; however, these events were absent among European-ancestry haplotypes. Two European haplotypes carrying *AACG* contained *CYP2D6–CYP2D7* fusion genes predicted to generate nonfunctional transcripts.
3. Among European-ancestry haplotypes, six of eight *AACG* haplotypes carried the rs3892097 T allele defining *CYP2D6*4*, whereas all *GGTA* haplotypes carried the reference C allele. Additional amino acid substitutions (P34S and S486T) were strongly correlated with *AACG* (Tables S7, S9). Analysis of phased haplotypes from the 1000 Genomes Project confirmed this enrichment: 631 of 642 *AACG* haplotypes carried at least one linked variant, including 231 carrying the rs3892097 splice-disrupting allele, whereas only 14 of 579 *GGTA* haplotypes carried any.
4. SNP array intensity values spanning *CYP2D6* were examined to assess linkage with CNV. Mean Log R Ratio values suggested increased copy number, with significant associations between intensity and *AACG* allele count, as well as a significant interaction with cocaine use frequency (Fig. 4; Table S11).

**Fig. 3.**
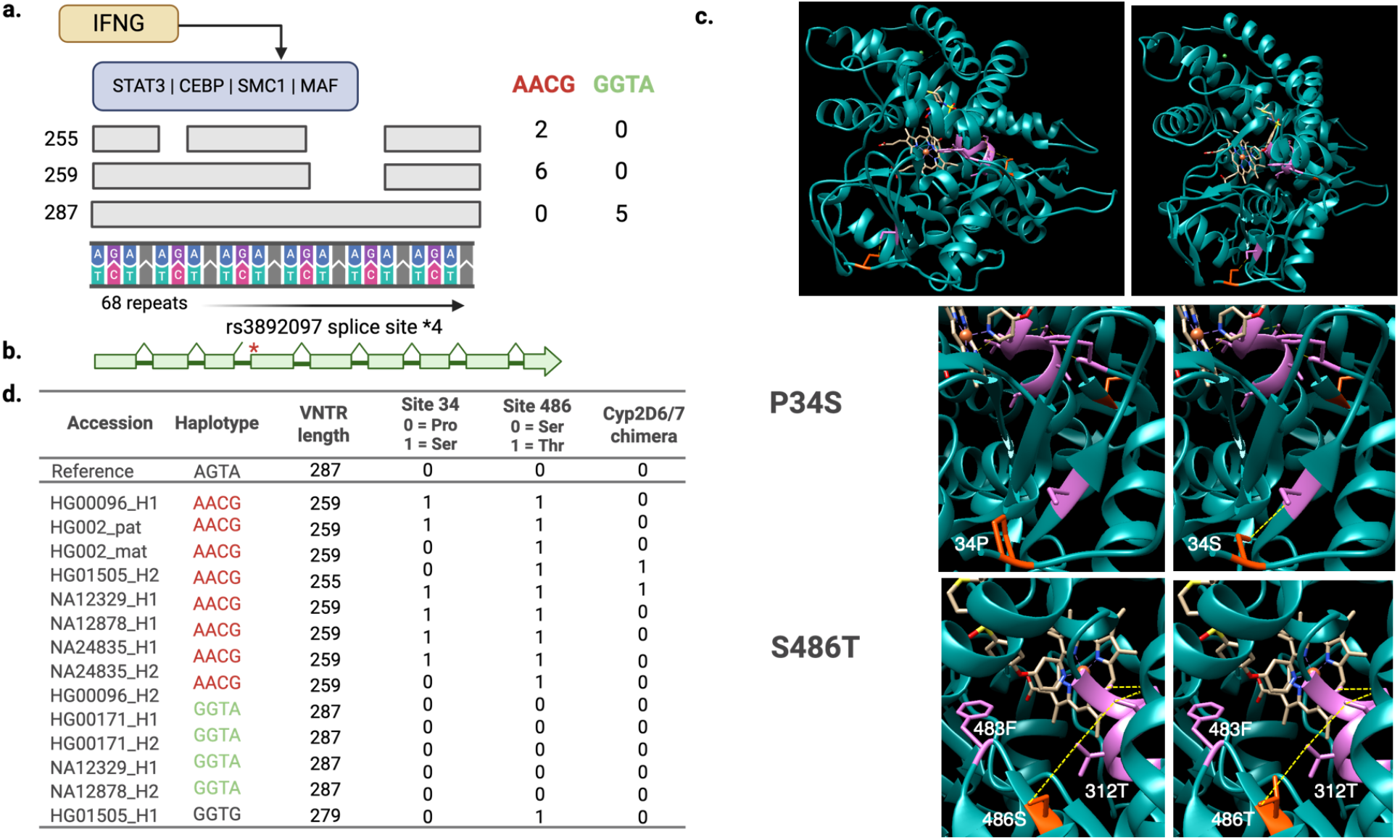
Features linked to CCC-derived alleles in the long-read haplotype assemblies. (a). The AACG risk allele is associated with the number of repeat units in the VNTR located in the NDUFA6-DT gene (AGAG, AGAC or AGAT). Deletions overlap a site for transcription factors that bind in response to interferon gamma (IFNG). All of those with the risk allele have deletions (gaps) but none of the individuals with the protective allele do. (b). SNP that defines the star 4 allele (no protein activity) in CYP2D6. A ‘T’ at this location disrupts a splice site just before exon 4 (red asterisk). (c). Three-dimensional structure of human CYP2D6 using PDB structure 2AM4 and chimera [35]. Substitutions P34S and S486T (orange residues) are located near the binding pocket and are predicted to alter hydrogen bonding (broken yellow lines) with residues in or near the binding pocket (pink residues). Edits were performed with the rotamer function and potentially interacting residues were those within six angstroms of -OH groups. (d). Table showing associations of the haplotypes from the long read 1000 Genome assemblies.

**Fig. 4.**
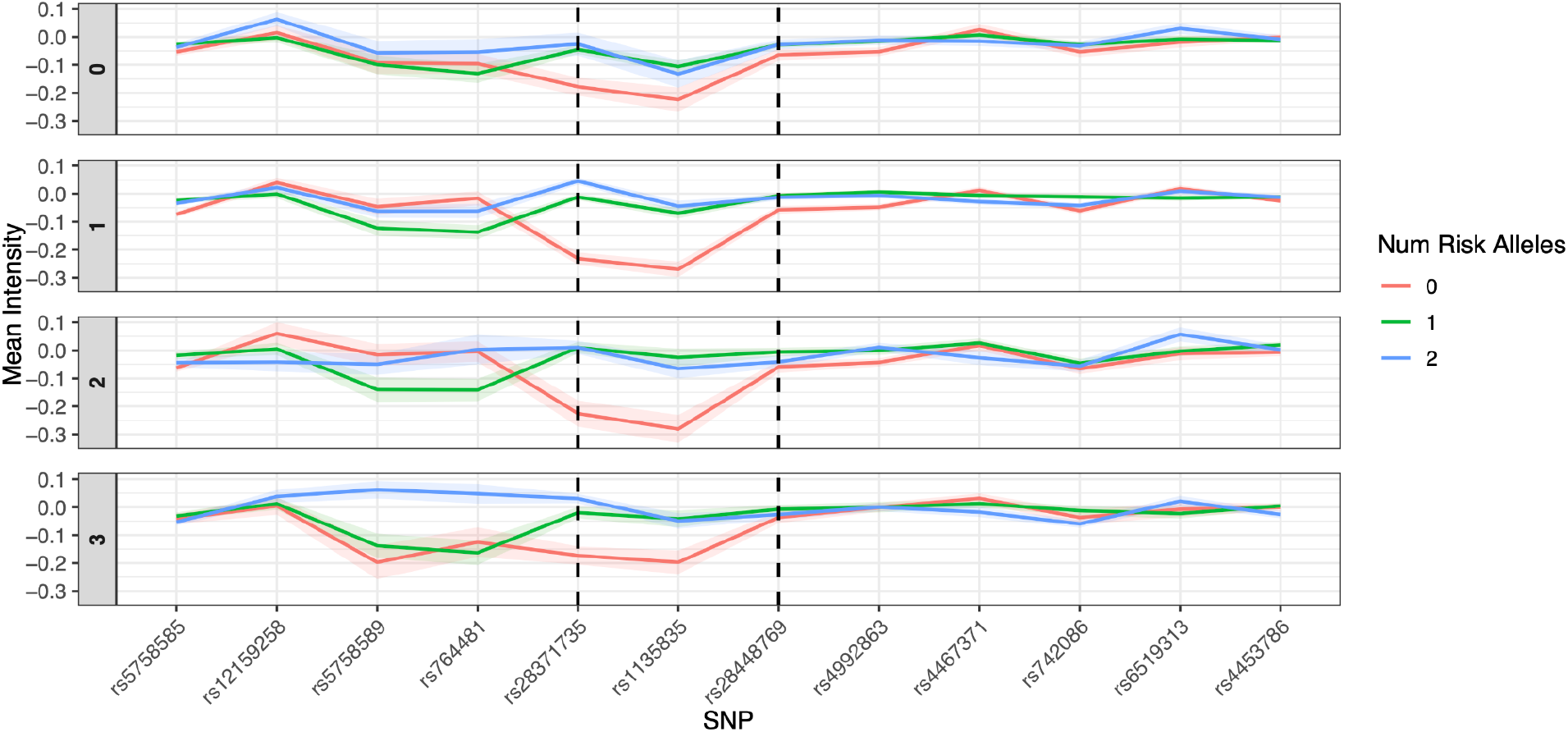
Plot of mean normalized intensity values (LogRR) of SNPs that cross the CYP2D6 locus for 2,155 individuals (blue rsIDs from Fig.2a). Gray boxes on the left of each panel indicate cocaine use frequency, broken line is the location of the CYP2D6 gene. Colored lines indicate the number of AACG risk alleles (0,1 or 2) and include uncertainty around the mean intensity values. Broken lines delineate the location of the CYP2D6 gene. A fixed effects glm determined a significant relationship between intensity values for SNPs rs5758589, rs764481, rs28371735, and rs1135835 and number of risk alleles as well as an interaction between risk allele and cocaine use (Table S11). For cocaine use 1 and 2, a difference in intensity is seen for SNPs rs28371735 and rs1135835 with carriage of at least one risk allele compared to no risk allele. For the highest cocaine users (bottom panel), individuals with two risk alleles (blue line) have increased intensity at SNPs rs5758589 and rs764481, indicating copy number increases in the genomic region in between NDUFA6-DT and CYP2D6.

Because the *AACG* haplotype is common in our cohort, these linked structural and sequence variants are likely segregating in the underlying population, providing a plausible genomic substrate for the observed regulatory and diplotype effects.

## Discussion

Here, we report the first allelic haplotypes significantly associated with cocaine use disorder that explicitly capture epistatic contributions to heritability. This success likely reflects the design of the eQTL-CCC framework, which identifies co-evolving regulatory variants and constructs multi-SNP haplotypes that jointly modulate gene expression. By preserving evolutionarily constrained variant combinations, this approach retains functional genetic structure that is often disrupted by standard SNP pruning strategies used to reduce computational burden or local linkage disequilibrium prior to association testing.

Notably, we were able to detect significant loci with relatively modest sample sizes, indicating that this framework may be particularly valuable for genetic studies in underrepresented populations where large cohorts are not yet available. Although we did not explicitly quantify the proportion of heritability attributable to epistasis, our results demonstrate that epistatic regulatory architectures represent a substantial and currently underappreciated component of genetics linked to addiction and substance abuse

### Cocaine use and HIV in African ancestry subpopulation

In support of this last point, we identified nine significant associations with cocaine use in subpopulation 4AFA, which is mainly of African ancestry. Several of the genes regulated by the eQTL haplotype networks are either directly involved in dopamine release or have demonstrated differential regulation upon administration of cocaine in animal models. In addition, six of the genes are linked to HIV infection and replication, providing potential insights into higher HIV rates in African Americans and increased infectivity in those who use cocaine [36,37]. Furthermore, allele haplotypes from two of the nine were statistically associated with HIV infection in the subgroup of individuals that speedball cocaine with heroin (inject both substances together). The deadly practice of combining opioids and stimulants has continued to the present [38] and resulted in more rapid increases in opioid/stimulant overdose mortality surpassed only by fentanyl associated overdose mortality [39], especially in the non-Hispanic African American population [40]. While non-Hispanic African American and Hispanic adults in the US living with HIV are much more likely to use cocaine than non-Hispanic White adults living with HIV [37], there was no significant association of any of the alleles in any of the ancestries with HIV overall, suggesting that this combination of drugs or their mechanism of administration (i.e. injection) may increase HIV infection by altering the expression of target genes regulated by the allele haplotype.

### Involvement of CYP2D6 in CUD in European ancestry subpopulation

The plot of intensity values from the SNP array indicates that our CCC-eQTL allele haplotype likely tags a structural variant (copy number increase) in the adjacent *NDUFA6-DT* and *CYP2D6* genes (Fig. 4). CYP2D6 is widely known in the pharmacogenomics field as it metabolizes nearly a quarter of pharmaceutical drugs [41,42]. From an evolutionary perspective, it likely evolved as an important metabolizer of exogenous xenobiotic compounds our ancestors ingested, including plant metabolites [43]. These two ideas are not incongruent as many of the recreational drugs ingested by humans, including cocaine, are secondary plant metabolites produced to combat herbivory [44]. What we have identified here may have evolved as a means of survival by communicating the toxicity of these compounds to the herbivore.

Although a major role of CYP2D6 is xenobiotic metabolism in the liver, it also plays an important role in normal brain function [45–48]. Furthermore, individuals with different haplotypes we identified here express different levels of *CYP2D6* in dopamine-producing areas of the brain, but do not differ in levels of expression in the liver. The enzyme does not metabolize cocaine and is in fact inhibited by it [49,50], but CYP2D6 can produce dopamine and serotonin through alternative pathways (Fig. 5,6) [24,47,48]. Genetic changes that alter CYP2D6 activity would theoretically have a direct effect on basal dopamine and serotonin levels and therefore our finding is consistent with its association with cocaine abuse as it has been well established that the immediate mechanism of action of cocaine is to block the reuptake of dopamine, serotonin and norepinephrine, increasing their half-life in the synapse [51–53]. Indeed, mice transgenic for human *CYP2D6* demonstrate elevated levels of serotonin in the brain [45].

**Fig. 5.**
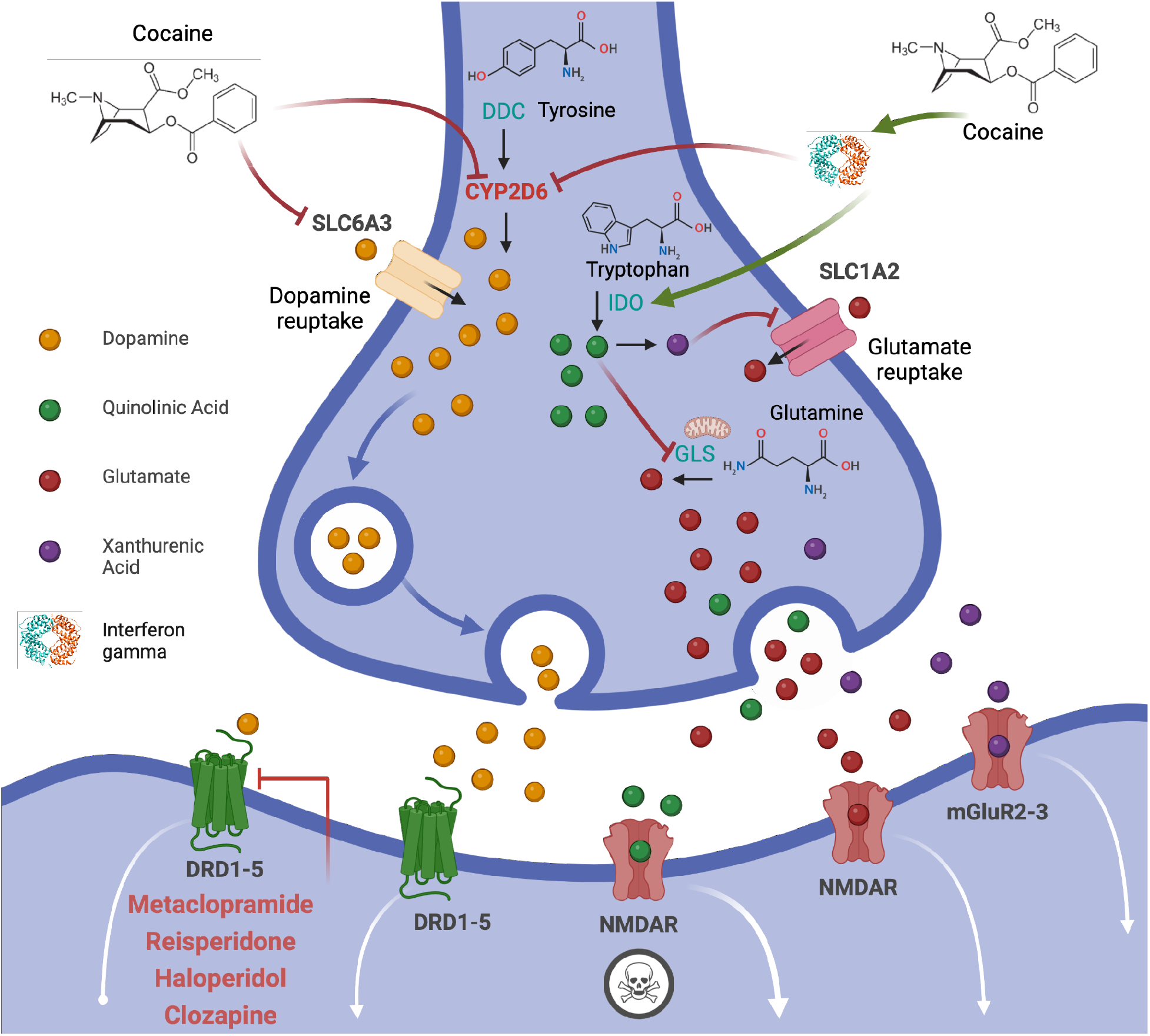
Model of a synapse as it relates to cocaine and CYP2D6. Short-term cocaine effects result from increases in dopamine in the synaptic cleft from blocking reuptake by SLC6A3. CYP2D6 can generate dopamine from tyrosine via an alternate pathway (normally from DOPA decarboxylase) and is inhibited by cocaine. Interferon gamma (IFNG) is induced by cocaine and increases expression of the IDO enzyme in the kynurenine pathway (also in Fig. 6), resulting in the production of the tryptophan metabolite quinolinic acid, which is toxic to NMDA receptors at high concentrations. The quinolinic acid derivative xanthurenic acid blocks the reuptake and synthesis of glutamate (the normal ligand for NMDA receptors) and is a ligand for metabotropic glutamate receptors (mGluRs). In addition to its role in catecholamine production, CYP2D6 breaks down dopamine receptor antagonists (e.g. neuroleptics) and therefore any changes to CYP2D6 function could affect dopamine levels. GLS = glutaminase, IDO = indoleamine 2,3-dioxygenase 1, DDC = dopa decarboxylase.

**Fig. 6.**
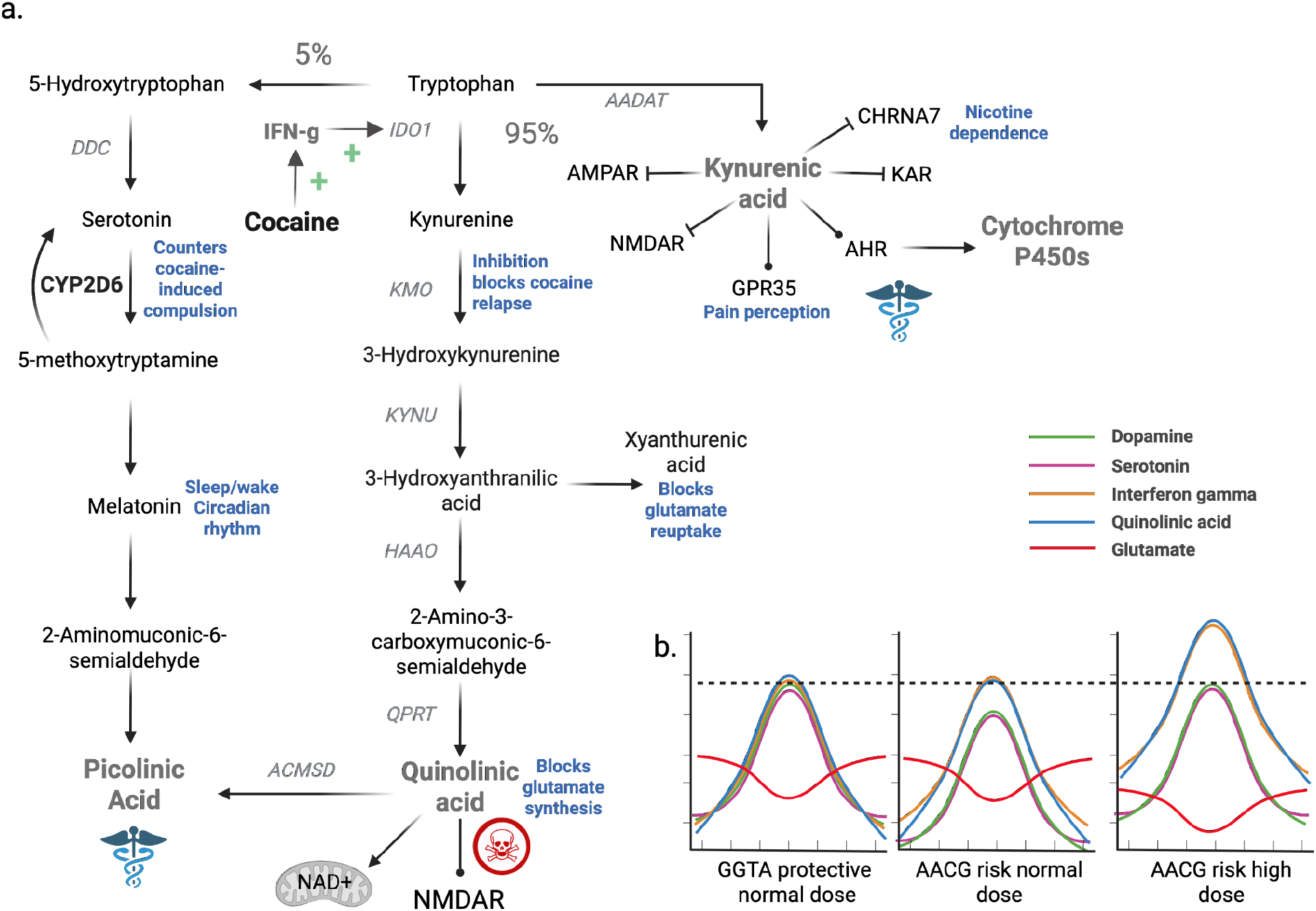
(a). CYP2D6 and the kynurenine pathway in response to cocaine. Tryptophan is the precursor for the metabolites in the pathway. Serotonin production accounts for ∼5% of the breakdown whereas the majority is shuttled to the generation of kynurenic (protective) and quinolinic (toxic) acids. CYP2D6 converts 5-methoxytryptamine to serotonin. Cocaine is a known inducer of interferon gamma, which activates IDO that converts tryptophan to kynurenine. Biological activities related to cocaine and features in Fig. 6 are in blue text. (b). Hypothetical model of the effect of cocaine administration in individuals with different alleles. In those with normal functioning CYP2D6 (left plot), cocaine induces a transient increase in dopamine, serotonin, interferon gamma, and quinolinic acid along with a decrease in glutamate. In individuals with the AACG risk allele (center plot), basal levels of dopamine and serotonin are lower, but the interferon gamma, quinolinic acid, and glutamate responses are similar to those with the protective GGTA allele. This induces these individuals to use cocaine more frequently or in greater amounts to achieve the same dopamine and serotonin levels (broken line), resulting in higher interferon gamma and quinolinic acid production, as well as reduced glutaminergic signaling (right plot), contributing to later stages of addiction. AADAT = aminoadipate aminotransferase, DDC = dopa decarboxylase, IDO = indoleamine 2,3-dioxygenase 1, KMO = kynurenine 3-monooxygenase, KYNU = kynureninase.

The *CYP2D6* locus is one of the most variable regions of the human genome with more than 100 alleles reported (pharmvar.org/gene/CYP2D6), but accurate genotypes are difficult to obtain given the highly repetitive and complex nature of this locus, and therefore the alleles are based on metabolism-based phenotypes rather than DNA-based genotypes [54]. These phenotype-based star alleles are generally binned into ultra, normal, intermediate, and poor metabolizers with the assumption that poor metabolizers have the gene deleted while increased metabolism is the result of gene duplications. Accurate genotyping is therefore of great importance when prescribing certain drugs. For example, codeine is metabolized to its active compound morphine by CYP2D6 and therefore current warning labels indicate that a poor-metabolizer would have no clinical benefit from administration of this drug, and an ultra-metabolizer may overdose on the standard prescribed amount (pharmgkb.org/gene/PA128).

Our analysis of 145 publicly available chromosome-level haplotype long read assemblies [55,56] indicates that the *AACG* risk haplotype is frequently associated with *CYP2D6* alleles predicted to be non-functional or severely altered, consistent with a poor-metabolizer phenotype. Among individuals of European ancestry, copy number gains of *CYP2D6* were observed only on *AACG* haplotypes, but all such copies are predicted to be non-functional. In addition, the majority of *AACG* haplotypes carry the T allele of rs3892097, which defines the *CYP2D6* *4 (“star 4”) haplotype and disrupts a splice acceptor site, resulting in a frameshifted transcript predicted to produce no functional protein (Table S4). Finally, all *AACG* haplotypes carry at least one amino acid substitution predicted to alter the ligand-binding pocket or active site of the enzyme.

Together, these results indicate that, in individuals of European ancestry, the *AACG* haplotype is strongly enriched for CYP2D6 protein with reduced or altered function, whereas the reference *GGTA* haplotype predominantly carries fully functional copies of the gene. This pattern is consistent with a diplotype-specific genetic architecture in which functional variation at CYP2D6 is determined by the combination of haplotypes carried by an individual, rather than by a simple dominant or recessive model.

Replication of the *CYP2D6* finding was performed in two polysubstance use datasets. The first dataset (phs000952.v1.p1) was collected to identify and characterize subtypes of and genetic loci contributing to cocaine and opioid use and dependence [57,58]. In the parent dataset phs000952.v1.p1, most individuals are cocaine and nicotine dependent, and slightly less than half are alcohol and opioid dependent [59]. In a sample from phs000952.v1.p1 with an effective sample size 43% of the discovery cohort, the risk diplotype (*AACG/AACG*) association was validated (*p*=0.017). The second dataset (phs001810.v2.p2) was collected to identify and characterize the genomics of crack cocaine use in polysubstance users; however, in phs001810.v2.p2, by definition, most cases are cocaine dependent, no controls are cocaine dependent, alcohol and nicotine dependence is common only in cases and opioid dependence is found in only a few cases. In a sample from phs001810.v2.p2 with an effective sample size 70% of the discovery cohort, the diplotype (*GGT/GGT*) had a protective effect of marginal significance; when the case:control sample was limited to ever smokers (effective sample size 25% of discovery cohort), the marginal protective diplotype effect remained but the risk diplotype effect increased (becoming similar to the effect observed in phs000952.v1.p1) and became marginally significant. These analyses in two different polysubstance datasets (the first with multiple drug dependencies common in both cases and controls, the second with cases cocaine dependent and mostly ever smokers and controls not cocaine dependent and mostly never smoking) supports our model of the *CYP2D6* risk diplotype having effects on tryptophan and serotonin metabolism in the presence of cocaine and nicotine. The observation that these drugs act at the large effect locus for cigarette smoking intensity and dependence [60,61] suggests genomic research should continue to examine the comorbid pair of cocaine and nicotine use and dependence, and other drug dependencies, associated with increased cardiovascular, respiratory and cancer mortality [62–64].

### Structural variation in the NDUFA6-DT, interferon gamma, and the kynurenine pathway

We found that the *AACG* risk allele is completely linked to the deletion of a VNTR unit in the 3’ end of the *NDUFA6-DT* gene that is a known binding site for transcription factors expressed in response to interferon gamma (Fig. 3, Table S5). More than two decades ago it was shown that interferon gamma is induced in response to cocaine administration [65]. Recent reports revealed that interferon gamma secreted in response to cocaine likely crosses the blood brain barrier via immune cells in the meningeal spaces [66].

Interferon gamma and CYP2D6 are also key components of the kynurenine pathway that can produce the monoaminergic neurotransmitter serotonin, picolinic acid, quinolinic acid, and kynurenic acid from the essential amino acid tryptophan depending upon which enzymes are active. The ratio of these compounds has been linked to numerous neuropsychiatric conditions including addiction [67]. In general, kynurenic and picolinic acid are neuroprotective and quinolinic is neurotoxic based on NMDA receptor-mediated excitotoxicity [68]. CYP2D6 can produce serotonin from the melatonin precursor 5-methoxytryptamine [69] and therefore, as is the case with dopamine, those with the *AACG* risk allele would be predicted to have reduced serotonin as a result of non-functioning CYP2D6. It has been widely reported that interferon gamma is an inducer of indoleamine 2,3-dioxygenase (IDO1) that converts tryptophan to kynurenine, which is eventually metabolized to quinolinic acid and NAD. Administration of cocaine tilts the metabolism of tryptophan toward toxic quinolinic acid and its derivatives and away from protective kynurenic acid.

Metabolites in the kynurenine pathway also directly affect the glutaminergic system, which has been implicated in CUD and other addiction disorders as an important component of the later stages of addiction [26,53]. Quinolinic acid is an agonist to NMDA receptors, causing hyperexcitability and eventual toxicity and also blocks the synthesis of glutamate from glutamine in the mitochondria (Fig. 5). In addition, a precursor to quinolinic acid, xanthurenic acid, blocks the reuptake of glutamate from the synaptic space [70] (Fig. 6a).

## Conclusions

### Summary and hypothesized model

We identified several allele haplotypes that are significantly associated with cocaine use in individuals of African American ancestry. Several of these are linked to HIV infection among individuals reporting combined heroin and cocaine use and may represent promising genetic links to the elevated rates of HIV infection observed among self-identified African American and Hispanic adults who use cocaine, where both mechanistic and social determinant–based translational research is warranted [37]. In addition, we identified a four-nucleotide haplotype that marks a locus associated with cocaine use and is strongly linked to structural and regulatory variation at *CYP2D6* in those of European ancestry.with heterozygotes carrying one copy of the *AACG* risk haplotype and one copy of the reference *GGTA* haplotype showing reduced cocaine use prevalence relative to either homozygous class

Phased diplotype analysis indicates that this locus exhibits an overdominant genetic architecture. This pattern suggests a heterozygote-advantage model in which the combination of haplotypes produces a more favorable neurochemical state than either homozygous configuration.

Long-read population assemblies indicate that the *AACG* haplotype is frequently associated with structural and sequence variants predicted to reduce or alter CYP2D6 function, including disruption of interferon gamma–responsive transcription factor binding sites, splice-disrupting alleles, and amino acid substitutions affecting the ligand-binding pocket. In contrast, the *GGTA* haplotype predominantly carries the reference functional form of the gene. A structural variant fully linked to the *AACG* haplotype disrupts an interferon gamma–responsive transcription factor binding site, providing a mechanistic link to the kynurenine pathway, which is known to play a role in cocaine use disorder.

Under this hypothetical model (Fig. 6b), individuals homozygous for the *GGTA* haplotype would be expected to exhibit baseline CYP2D6 activity, such that cocaine exposure induces transient increases in dopamine, serotonin, interferon gamma, and quinolinic pathway metabolites, consistent with observations in the humanized CYP2D6 transgenic mouse [45] and a corresponding reduction in glutamate synthesis. In contrast, individuals homozygous for the *AACG* haplotype would be expected to exhibit reduced or altered CYP2D6 function, leading to lower basal levels of dopamine and serotonin and requiring higher or more frequent cocaine dosing to achieve equivalent neurochemical reinforcement. This, in turn, would be predicted to amplify interferon gamma signaling and activation of the kynurenine pathway, disrupting glutamatergic signaling via impaired glutamate synthesis, reduced reuptake, and preferential activation of metabotropic glutamate receptors rather than NMDA receptors.

Heterozygotes carrying one copy of each haplotype may occupy an intermediate and more stable neurochemical state, in which basal neurotransmitter levels and cocaine-induced perturbations are buffered relative to either homozygous class. This configuration may represent an optimal balance of dopaminergic, serotonergic, and glutamatergic signaling, providing a plausible molecular basis for the observed heterozygote advantage.

### Limitations and future directions

Portions of our proposed model of *CYP2D6* dysfunction are based on publicly available long-read genome assemblies from individuals without known cocaine use. Further validation will require direct long-read sequencing of *CYP2D6* in individuals with documented cocaine use to confirm the inferred structural variation and copy number changes. In addition, our findings are based on a cohort consisting primarily of opioid users with a subset of cocaine users, which may limit generalizability. However, polysubstance use is common among individuals who use illicit drugs. Future studies in independent cocaine-use cohorts will be necessary to replicate these associations and refine the contribution of epistatic regulatory architectures to addiction and substance abuse risk. Expected case counts were derived from the haplo-diplotype distribution among unaffected participants. This approach treats the unaffected sample frequencies as estimates of the underlying reference distribution and does not incorporate uncertainty in those estimates; therefore, significance may be overstated when the control sample or an individual haplo-diplotype category is small.

## Methods

### Study Subjects

The subjects in these analyses were enrolled in the Weiss prospective cohort study, a set of related projects initiated by SH Weiss in the 1980’s. All subjects had given written consent. Only those with a history of opioid use disorder were selected for inclusion [71–76]. All were interviewed in person about their use of cocaine in the prior five years, and their frequency of cocaine use in the prior year. Those who had used cocaine in the prior five years but did not provide a frequency of use in the past year were treated as missing frequency of cocaine use, whereas those who had not used cocaine at all in the prior five years were treated as non-users. Additional extensive information about other substances and factors has been collected. For those interviewed more than once, their first complete interview data were used. The study was reviewed and approved by the Rutgers Institutional Review Board. For replication, we used two cohorts cohort available through the NIH dbGAP portal (Substance Dependence GWAS in European- and African - Americans, dbGaP Study Accession, phs000952.v1.p1 and Gene-Environment Interactions in COCCaINE Use Disorder: Collaborative Case-Control Initiative in Cocaine Addiction, phs001810.v2.p2).

### Genome Wide Association Study

Although a GWAS was not the focus of this study, we performed the analysis as a contrast to our epistasis approach. DNA extraction, amplification and genotyping were done in conjunction with a NIDA contracted lab in accordance with NIDA protocols [77]. We genotyped 9,725 unrelated individuals at 714,238 loci on the Illumina OmniExpress array (Fig.1). We then filtered for individuals with a cocaine use phenotype, a call rate of > 0.85 leaving 8,481 samples for analysis. We then removed duplicate samples (some individuals had more than one sample genotyped) and we also removed samples from related individuals based on the kinship function in PLINK2. In these cases, we retained the individual with the highest call rate leaving 8,029 individuals.

To account for population substructure, we used the ancestry population groups determined by GRAF-POP [18]. The sample sizes for four of the groups were small (2AFR = 91, 3EAS = 9, 7OAS = 16, 9OTH = 87) and therefore we restricted out analyses to the other four groups (1CEU = 2,699, 4AFA = 3,008, 5HS1 = 552, and 6HS2 = 1,567) for a total of 7,826 individuals. Self-reported race is provided, but all analyses used genetic-based subpopulations to account for substructure.

A genome wide association study was performed for each of the four populations (1CEU, 4AFA, 5HS1, and 6HS2) using the “glm” model in PLINK2 for the phenotype of cocaine use that included frequency of use, coded as follows:

1. no use in the prior five years;
2. use less than once per week;
3. use at least once per week but less than once per day; and
4. use at least once per day.

The first ten components of a PCA and sex were used as covariates to account for any remaining population substructure. The PCA was generated using randomly pruned SNPs in PLINK for each of the four populations separately with the indep-pairwise function (window size = 1000, step size = 50, *r*^2^ = 0.3). Cocaine use, coded as a binary outcome, was analyzed using logistic regression with PLINK’s logistic–Firth hybrid procedure, which applies Firth penalized regression when standard logistic regression encounters separation or convergence problems. Frequency of cocaine use was analyzed as a quantitative outcome using linear regression. We utilized a Bonferroni correction within each population GWAS for genome-wide significance, and a second threshold of 1 × 10^-5^ to designate suggestive genome-wide significance.

### Epistasis/BlocBuster on eQTLs

We wished to reduce multiple testing burdens typically seen with GWAS by jointly testing sets of SNPs that potentially represent epistasis. There are many ways to define potentially relevant SNP sets (e.g. group all SNPs within each gene). Here we chose to focus on sets of eQTLs with potential joint regulatory effects on cocaine use. We subset the array SNPs to only those listed as expression quantitative trait loci (eQTL) by the Genotype-Tissue Expression (GTEx, version 8) in all tissues and theORegAnno databases [78], leaving 49,393 SNPs. We then used BlocBuster’s CCC metric to calculate pairwise correlations of all 98,746 alleles (SNPs were bi-allelic) [14].

CCC performs correlation between a pair of biallelic SNPs *i* and *j* with alleles A/a and B/b respectively. Unlike other SNP correlation measures (e.g. *r*^2^ of PCC), CCC produces a value for each possible allelic combination for *i* and *j* (AB, Ab, aB, ab). If a pair of alleles (e.g. SNP*i*_a and SNP*j*_B) are found in combination at greater frequency than expected by chance, then CCC may return a high value specifically for that allelic combination, even if other allelic combinations at SNPs *i* and *j* are uncorrelated.

In previous work we found that a 0.7 CCC threshold produces a stringent allele correlation network that also reduces spurious correlations [79]. With smaller sample sizes, this correlation threshold can introduce noise and therefore for each subpopulation, we generated a null data set by randomly permuting genotypes among individuals and increasing the CCC threshold from 0.7 until no correlations were found (1CEU = 0.70, 4AFA = 0.75, 5HS1 = 0.85, 6HS2 = 0.8). We also filtered SNPs with a MAF ≥ 0.05 and those missing in more than 10% of individuals. These filters reflect a methodological choice aligned with haplotype-based, interaction-focused analyses rather than a constraint imposed by SNP density or genotyping platform. Correlations above the cutoff were then clustered by BlocBuster using breadth-first-search algorithm to identify eQTL allelic subnetworks within the population. Each of these subnetworks represents a multi-locus eQTL epistatic haplotype, which itself may have multiple alleles in the subpopulations. For example, three distinct eQTLs with alleles A/C, G/T and A/G can form a 3-locus CCC haplotype with alleles such as ATG, CTG, AGA etc. BlocBuster identifies the primary multi-locus allele that drives the formation of each subnetwork based on the frequency in the entire sample (i.e. cases and controls combined).

Counts of the allele networks (multi-locus haplotypes) were then generated with the same classification we used for the GWAS, but the analysis used a binary system where cases were those with a cocaine use frequency 1, 2, or 3 and controls = 0, or no use. This was done for the same six subpopulations as the GWAS. Allele networks were tested for significant association with cocaine use using a *chi*-square test followed by Bonferroni correction for multiple tests based on the number of allele haplotype networks. It is important to note that the CCC algorithm will include haplotypes that represent random linkage disequilibrium (LD), but these would be highly unlikely to show significant association with cocaine use because LD should be randomly distributed between case and control groups.

### Counts of allele eQTL haplotypes within different genotypes

For networks with significant associations, phased multi-locus SNP genotypes (termed haplo-diplotypes) were determined for each individual within the corresponding subpopulation. For individuals homozygous for the entire eQTL allele haplotype, or heterozygous at a single site, there is only one possible phased outcome. For individuals with more than one heterozygous site in the haplotype, the first allele was assumed to be the CCC-derived allele and the remaining nucleotides were used to build the second allele. These are identified with a slash rather than a pipe as they are predicted.

A *chi*-square test was then performed for each phased diplotype against cocaine use to determine if there were classic dominant/recessive patterns or non-additive effects of the CCC-allele. As was the case with the CCC presence/abscence allele, the frequency of each haplo-diplotype in the controls was used to calculate the expected number of that diplotype in cases. Phased diplotypes with fewer than 5 individuals were removed from the analysis due to the unreliability of the *chi*-square test below these values. A subset of the CCC-positive networks altered the expression of genes linked to HIV biology and therefore we performed a statistical test with the phased diplotypes for HIV infection. We also did this within only those that speedball (use heroin and cocaine together through injection) because of the higher HIV rate in that population.

For the single tissue GTEx analysis linked to the eQTL haplotype that includes CYP2D6, we averaged the normalized effect size (NES) for all tissues across the SNPs rs133335, rs7245, rs2284087, and rs5758589 (Table S5). If there was no reported effect for a SNP in a tissue for a gene (e.g. liver), it was reported as zero.

### Independent replication using the Yale-Penn dbGaP cohort

We evaluated the original multilocus findings in an independent substance-dependence cohort obtained through the NIH Database of Genotypes and Phenotypes (dbGaP accession phs000952.v1.p1; Substance Dependence GWAS in European- and African-Americans) [57,58]. Genotype data were generated using the Illumina HumanOmni1-Quad v1.0 array and were provided in PLINK binary format. Phenotypes were derived from the Semi-Structured Assessment for Drug Dependence and Alcoholism (SSADDA), with cocaine dependence defined according to DSM-IV criteria. Participants coded as cocaine dependent (‘cocdep = 2’) or unaffected by cocaine dependence (‘cocdep = 1’) were retained; participants coded as having cocaine abuse (‘cocdep = 0’) or with missing phenotype data were excluded. Because the deposited phenotype table did not contain a separate measure of lifetime cocaine exposure, participants coded as unaffected were considered free of a cocaine-dependence diagnosis but could not be confirmed to be cocaine-non-exposed controls.

The primary replication analysis was restricted to Caucasian non-Hispanic participants (‘A8a_RACE = 6’). Relatedness was evaluated from the genome-wide directly genotyped markers using PLINK 2.0 and the KING-robust estimator. To obtain a sample of unrelated individuals, one member of each pair with an estimated kinship coefficient greater than 0.0884 was removed using the PLINK “--king-cutoff 0.0884” procedure. This threshold excludes second-degree or more closely related pairs. Relatedness filtering was performed using genome-wide markers before extraction of the loci of interest. The resulting analysis included 892 unrelated participants, comprising 708 individuals with cocaine dependence and 184 individuals without a cocaine-dependence diagnosis.

The prespecified SNPs defining the original multilocus findings were extracted directly from the array genotypes without imputation. The complete SNP sets for Blocs 3480 and 3110 were unavailable (Table S4) and therefore we only validated Bloc 5500 (the *CYP2D6* haplotype). Alleles were harmonized to the strand and SNP order used in the discovery cohort, including strand complementation where required. As with the Weiss cohort, individual multilocus genotype strings were constructed by concatenating the two called alleles at each SNP in genomic order. Haplo-diplotypes with fewer than 10 individuals were removed and expected numbers of cases were derived from the frequency of controls. A chi-square test with Bonferroni correction was used to test for significance. Analyses were restricted to directly genotyped variants from the Illumina HumanOmni1-Quad v1.0 array; imputed variants were not used.

### Independent analysis of the COCCaINE cohort

An independent analysis was performed using controlled-access genotype and phenotype data from the COCCaINE study obtained through the NIH Database of Genotypes and Phenotypes (phs001810.v2.p2). Participants were genotyped using the Axiom Precision Medicine Research Array–SmokeScreen array, which contained 532,839 directly genotyped variants. Imputed genotypes were not used.

The phenotype table contained 1,874 participants and included self-reported race, lifetime cocaine use, cocaine abuse, cocaine dependence and other substance-use variables.. Participants with ever_co = 5 were classified as lifetime cocaine users (cases), whereas those with ever_co = 1 were classified as cocaine never-users (controls). Opioid use was an exclusion characteristic of this cohort and was rare in the deposited data; therefore, no opioid-based restriction was applied.

Relatedness was estimated from the genome-wide directly genotyped markers using the KING-robust estimator as was implemented in the Yale-Penn cohort. The discovery analysis defined the *CYP2D6*-associated haplotypes using four SNPs: rs133335, rs7245, rs2284087 and rs5758589. Because the COCCaINE PLINK matrix used Axiom probe identifiers rather than rsIDs, markers were identified by their GRCh37 genomic coordinates. Three markers were directly represented: rs133335 by AX-11250485 at 22:42,416,056; rs7245 by AX-40924061 at 22:42,481,849; and rs2284087 by AX-40924065 at 22:42,485,671. The fourth marker, rs5758589 was absent from the quality-controlled matrix and therefore the analysis evaluated a prespecified three-SNP proxy rather than the complete four-SNP haplotype. No neighboring marker was substituted for the missing variant, and no imputation was performed.

Alleles were harmonized to the strand and order used in the discovery cohort. Removal of rs5758589 reduced the principal discovery haplotypes *AACG* and *GGTA* to the three-marker proxies *AAC* and *GGT*, respectively. The final analysis included 1,057 unrelated White participants with complete three-SNP genotypes and cocaine-use phenotypes: 671 cocaine never-users and 386 lifetime cocaine users. After removing haplo-diplotypes with less than 10 individuals there were 652 controls and 366 cases. In addition, we analyzed only those that were smokers, to more closely resemble the polysubstance abuse individuals in the Weiss cohort, leaving 115 controls and 235 cases.

### Repeated subsampling analysis of the CYP2D6 association

To assess the robustness of the *CYP2D6* finding within the Weiss cohort, we performed a repeated random subsampling analysis. The cohort was randomly partitioned into 70% discovery and 30% validation subsets using 100 independent random seeds. For each partition, the CCC analysis was repeated using the same analytical parameters applied in the primary analysis. We then determined whether the previously identified *CYP2D6* signal containing the *GGTA* haplotype was recovered and recorded its estimated effect size and statistical significance. This procedure was intended as a within-cohort stability analysis rather than as an independent replication, because individuals necessarily occurred in multiple random partitions.

### Structural variation prediction and analysis at CYP2D6

Genomic structural variants can be inferred from the intensity values generated by Illumina OmniExpress arrays [80]. Intensity files were available for all samples, 2208 of which were from subpopulation 0 and had full genotypes for all four SNPs from the *CYP2D6* CCC haplotype. We extracted the LogRR values for the SNPs covering *CYP2D6*, *CYP2D7*, and *CYP2D8P* (rs5758585, rs12159258, rs5758589, rs764481, rs28371735, rs1135835, rs28448769, rs4992863, rs4467371, rs742086, rs6519313, and rs4453786). Each sample was categorized as having 0, 1, or 2 copies of the *AACG* risk allele based on the CCC eQTL SNPs (rs7245, rs2284087, and rs5758589). Statistical significance of intensity values with number of risk alleles and cocaine use was determined with a linear fixed-effects model in the *lme4* package in R.

## Data Availability

All data use to produce these results are available online in the NIH database of genotypes and phenotypes.

https://dbgap.ncbi.nlm.nih.gov/home/

## Declarations

### Ethics approval and consent to participate

This research was conducted in line with the principles of the Declaration of Helsinki. Approval was granted by the Rutgers Institutional Review Board (IRB) under protocol Pro20150001314, this protocol initially approved on 17 June 2015 and with continuing reviews approved each year since then. The replication analyses involved secondary analysis of de-identified data obtained from the database of Genotypes and Phenotypes (dbGaP) under authorized data-use agreements. Participants in the contributing dbGaP studies provided informed consent under protocols approved by their respective IRBs. The secondary analyses conducted for the present study were determined that this protocol is exempt from the federal regulations governing protection of human research participants under 45 CFR 46.104(d) by the Oregon Research Institute IRB.

### Funding/Acknowledgements

Research reported in this publication was supported by the National Institute on Drug Abuse (NIDA) of the National Institutes of Health under Award Numbers R01DA050495, as well as the Laboratory Directed Research & Development Program of Oak Ridge National Laboratory, managed by UT-Battelle, for the US Department of Energy (DOE). Also supported by the New Jersey Commission on Cancer Research under grant number COCR26PPR013. Biospecimens were genotyped under NIDA Contract N01DA187789. The content is solely the responsibility of the authors and does not necessarily represent the official views of the National Institutes of Health. This research used resources from the Knowledge Discovery Infrastructure at the Oak Ridge National Laboratory, which is supported by the Office of Science of the U.S. Department of Energy under Contract No. DE-AC05-00OR22725. Figures 5 and 6 were made with BioRender.

We acknowledge the Principal Investigator, Joel Gelernter, MD, Yale University School of Medicine, and the Co-Principal Investigator, Henry R. Kranzler, MD, University of Pennsylvania School of Medicine, Submitting Investigators who submitted data [5,81], and dbGaP for accession phs000952.v1.p1, and the National Institute on Drug Abuse and National Institute on Alcohol Abuse and Alcoholism, National Institutes of Health for funding RC2 DA028909, R01 DA12690, R01 DA12849, R01 DA18432, R01 AA11330, R01 AA017535, and for the VA Connecticut and Philadelphia VA MIRECCs. Part of the genotyping was supported by the Center for Inherited Disease Research (CIDR) and Yale University (Center for Genome Analysis). CIDR is fully funded through a federal contract from the National Institutes of Health to The Johns Hopkins University (contract number N01-HG-65403).

Funding support for the Genetic and Environmental Risk Factors for Cocaine Addiction - COllaborative Case-Control INitiativE in crack addiction – (COCCAINE) study (phs001810.v2.p2) was provided by the CNPq, CAPES, FAPERGS and SENAD through individual grants of Prof. Dr. Rodrigo Grassi-Oliveira, Prof. Dr. Claiton Bau and Prof. Dr. Flávio Pechansky. Data for the COCCAINE project were provided by Prof. Dr. Rodrigo Grassi-Oliveira at Pontifical Catholic University of Rio Grande do Sul, and Prof. Dr. Felix Kessler, Prof. Dr. Lisia Von Diemen and Prof. Dr. Eugênio Grevet at Federal University of Rio Grande do Sul. Assistance with phenotype harmonization and DNA extraction was provided by Dr. Rodrigo Grassi-Oliveira, Dr. Diego Rovaris and Dr. Breno Sanvicente-Vieira. The DNA was genotyped on the Smokescreen Chip by Rutgers University Cell and DNA repository supported by contract N01DA-14-7789.

### Author Contributions

MG – Conceptualization, Methodology, Validation, Formal Analysis, Investigation, Writing the Original Draft, Review and Editing, Visualization, Funding Acquisition

DR – Validation, Resources, Data Curation, Review and Editing, Project administration

AB – Conceptualization, Validation, Investigation, Review and Editing

DK – Conceptualization, Methodology, Validation, Writing Original Draft, Review and Editing, Visualization

SC – Conceptualization, Methodology, Software, Review and Editing

PA – Resources, Data Curation, Review and Editing

SP – Formal analysis, Data Curation

QZ – Formal analysis, Data Curation

SW – Conceptualization, Methodology, Investigation, Resources, Data Curation, Review and Editing, Supervision, Project Administration, Funding Acquisition

## Supplementary Material

## Supplementary Material

**Fig. S1.**
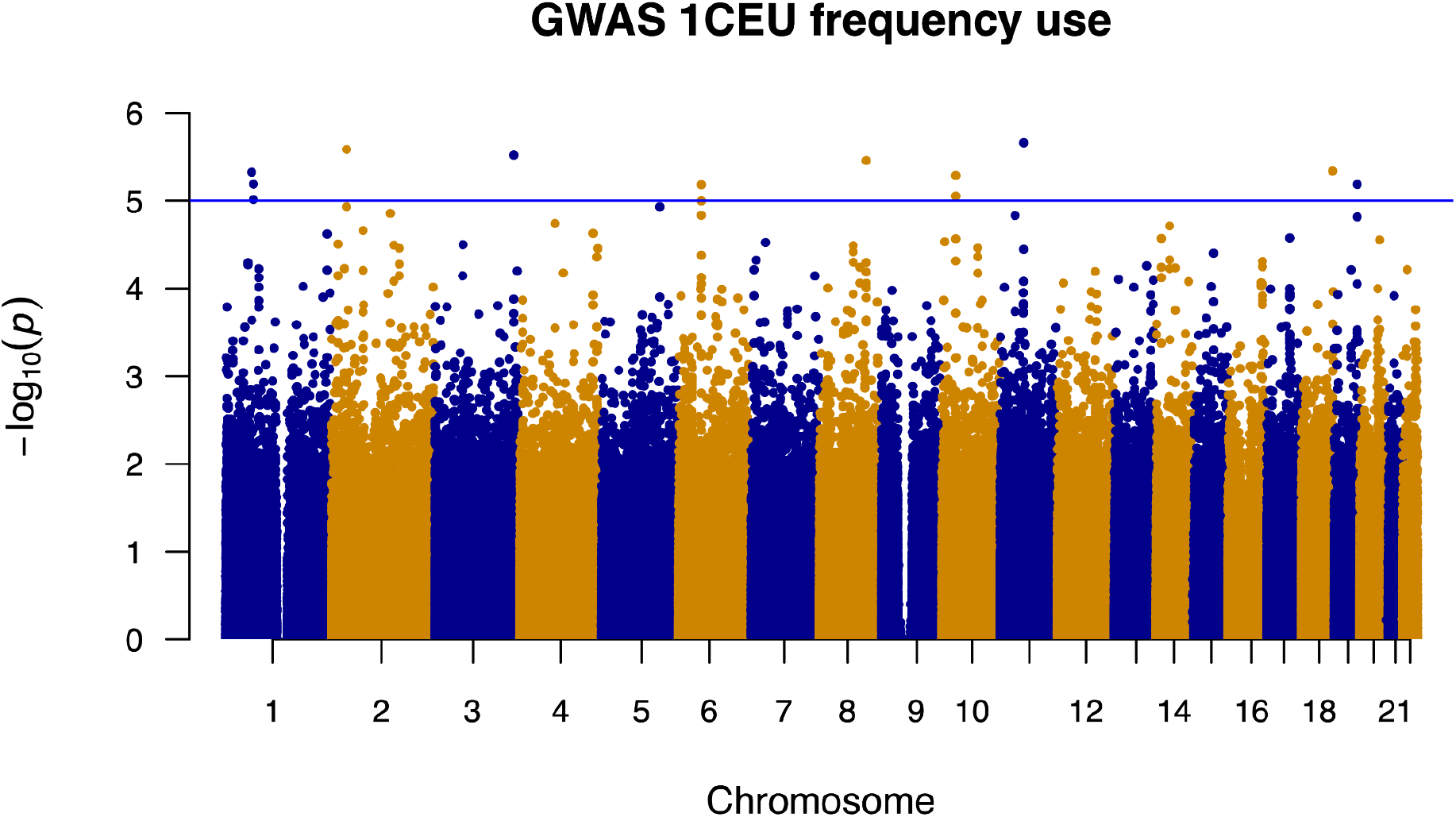
GWAS results for cocaine use frequency for (S1) 1CEU, mainly European ancestry, (S2) 4AFA, mainly African American ancestry, (S3) 5SH1, mainly Puerto Rican Hispanic ancestry, (S4) 6HS2, mainly Central and South American Hispanic ancestry. In each plot, the red line indicates statistical significance after Bonferroni correction (p < 5.5 10^-8^), blue line indicates suggestive significance (p < 1 × 10^-5^).

**Fig. S2.**
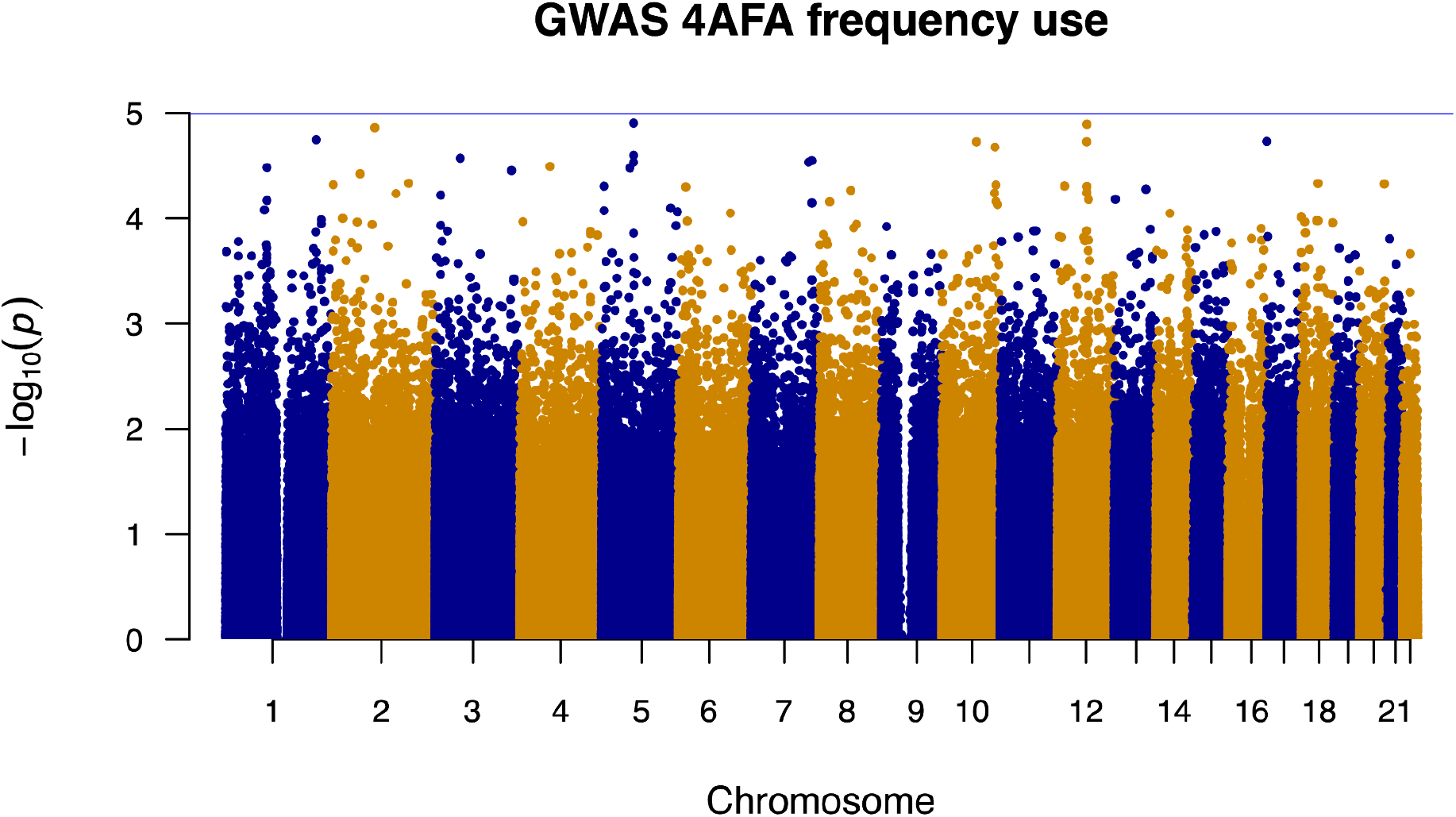
GWAS results for cocaine use frequency for (S1) 1CEU, mainly European ancestry, (S2) 4AFA, mainly African American ancestry, (S3) 5SH1, mainly Puerto Rican Hispanic ancestry, (S4) 6HS2, mainly Central and South American Hispanic ancestry. In each plot, the red line indicates statistical significance after Bonferroni correction (p < 5.5 10^-8^), blue line indicates suggestive significance (p < 1 × 10^-5^).

**Fig. S3.**
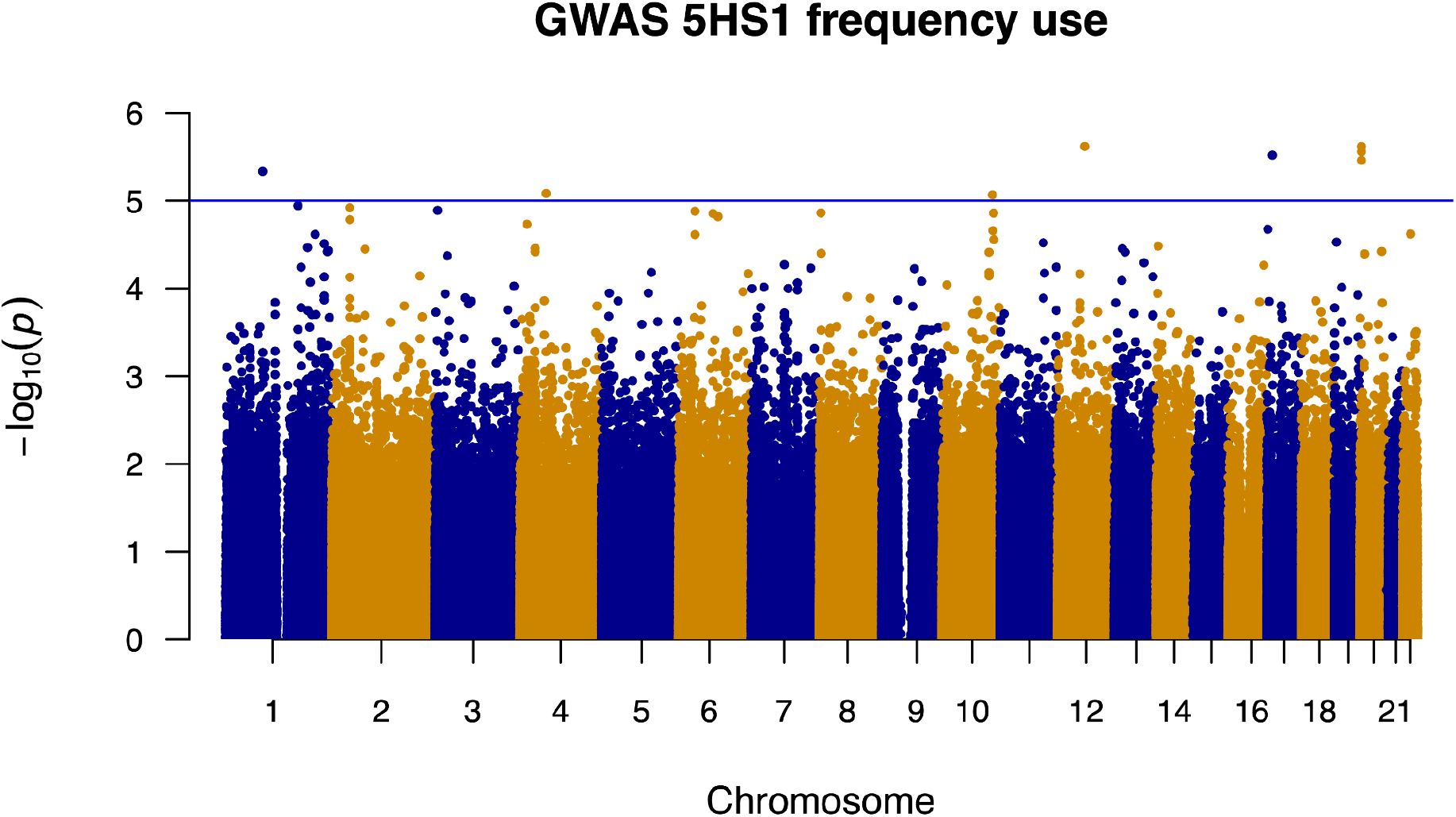
GWAS results for cocaine use frequency for (S1) 1CEU, mainly European ancestry, (S2) 4AFA, mainly African American ancestry, (S3) 5SH1, mainly Puerto Rican Hispanic ancestry, (S4) 6HS2, mainly Central and South American Hispanic ancestry. In each plot, the red line indicates statistical significance after Bonferroni correction (p < 5.5 10^-8^), blue line indicates suggestive significance (p < 1 × 10^-5^).

**Fig. S4.**
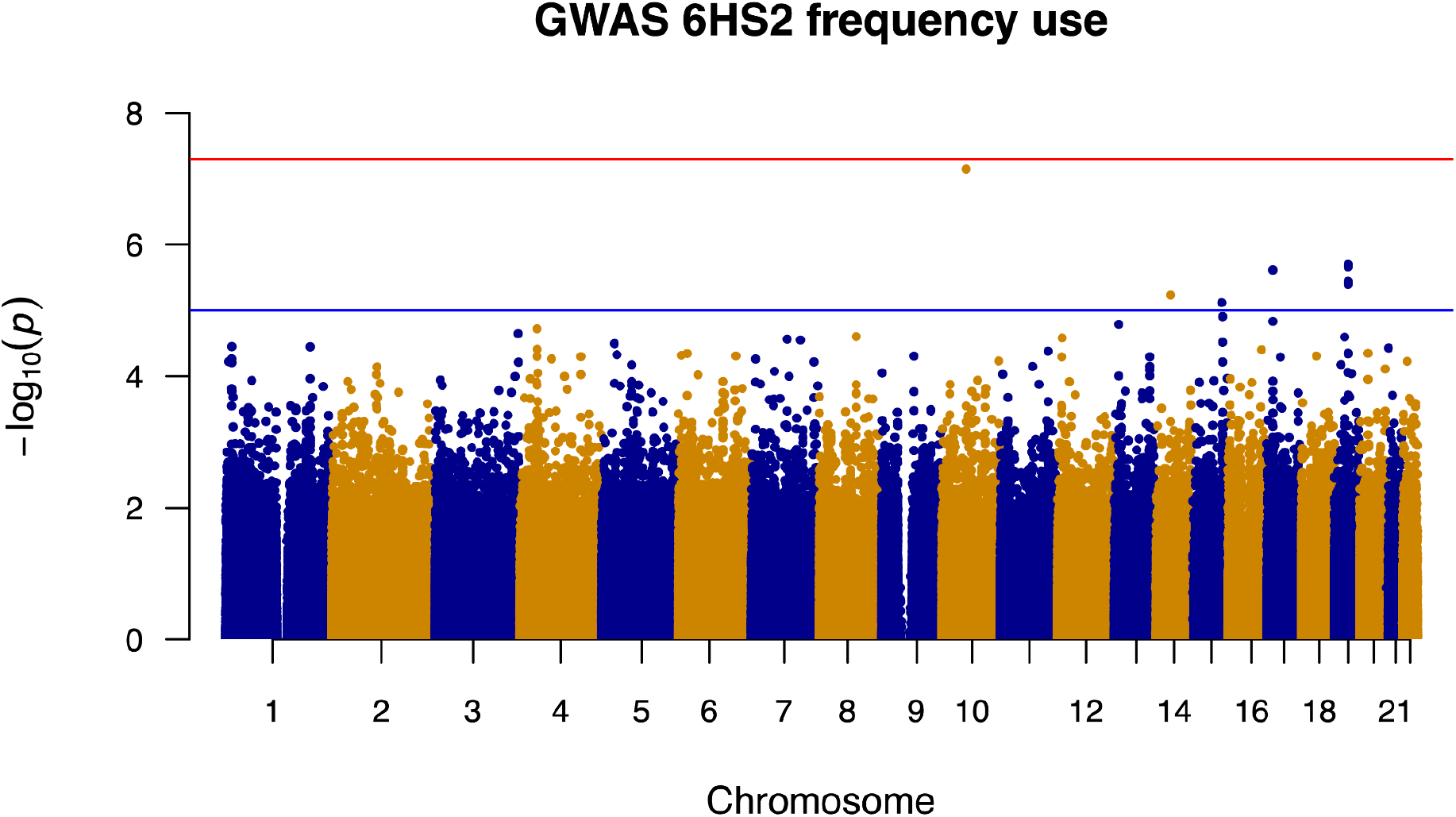
GWAS results for cocaine use frequency for (S1) 1CEU, mainly European ancestry, (S2) 4AFA, mainly African American ancestry, (S3) 5SH1, mainly Puerto Rican Hispanic ancestry, (S4) 6HS2, mainly Central and South American Hispanic ancestry. In each plot, the red line indicates statistical significance after Bonferroni correction (p < 5.5 10^-8^), blue line indicates suggestive significance (p < 1 × 10^-5^).

**Fig. S5.**
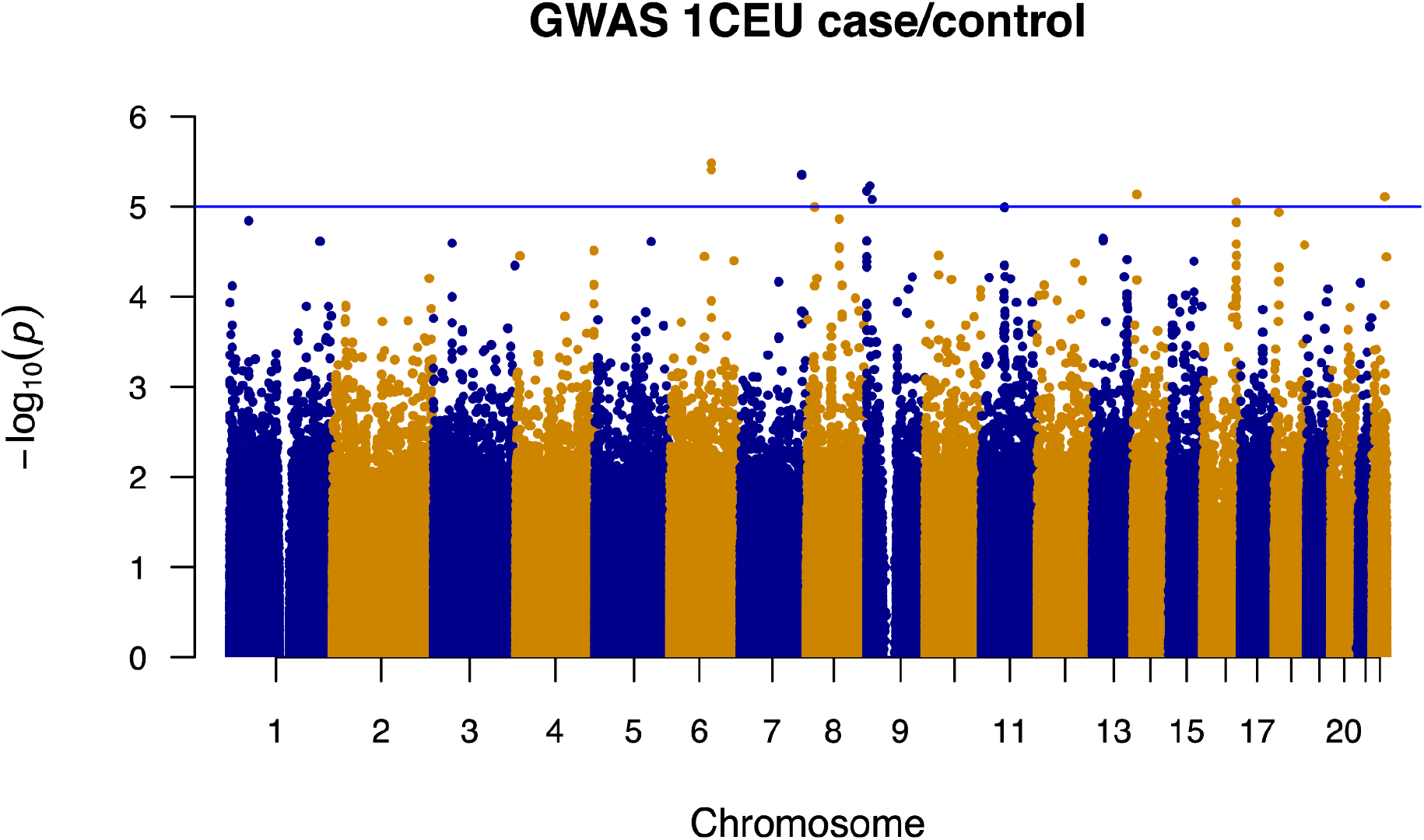
Fig. S1-S4. GWAS results for cocaine use (case) or no use (control) for (S1) 1CEU, mainly European ancestry, (S2) 4AFA, mainly African American ancestry, (S3) 5SH1, mainly Puerto Rican Hispanic ancestry, (S4) 6HS2, mainly Central and South American Hispanic ancestry. In each plot, the red line indicates statistical significance after Bonferroni correction (p < 5.5 10^-8^), blue line indicates suggestive significance (p < 1 × 10^-5^).

**Fig. S6.**
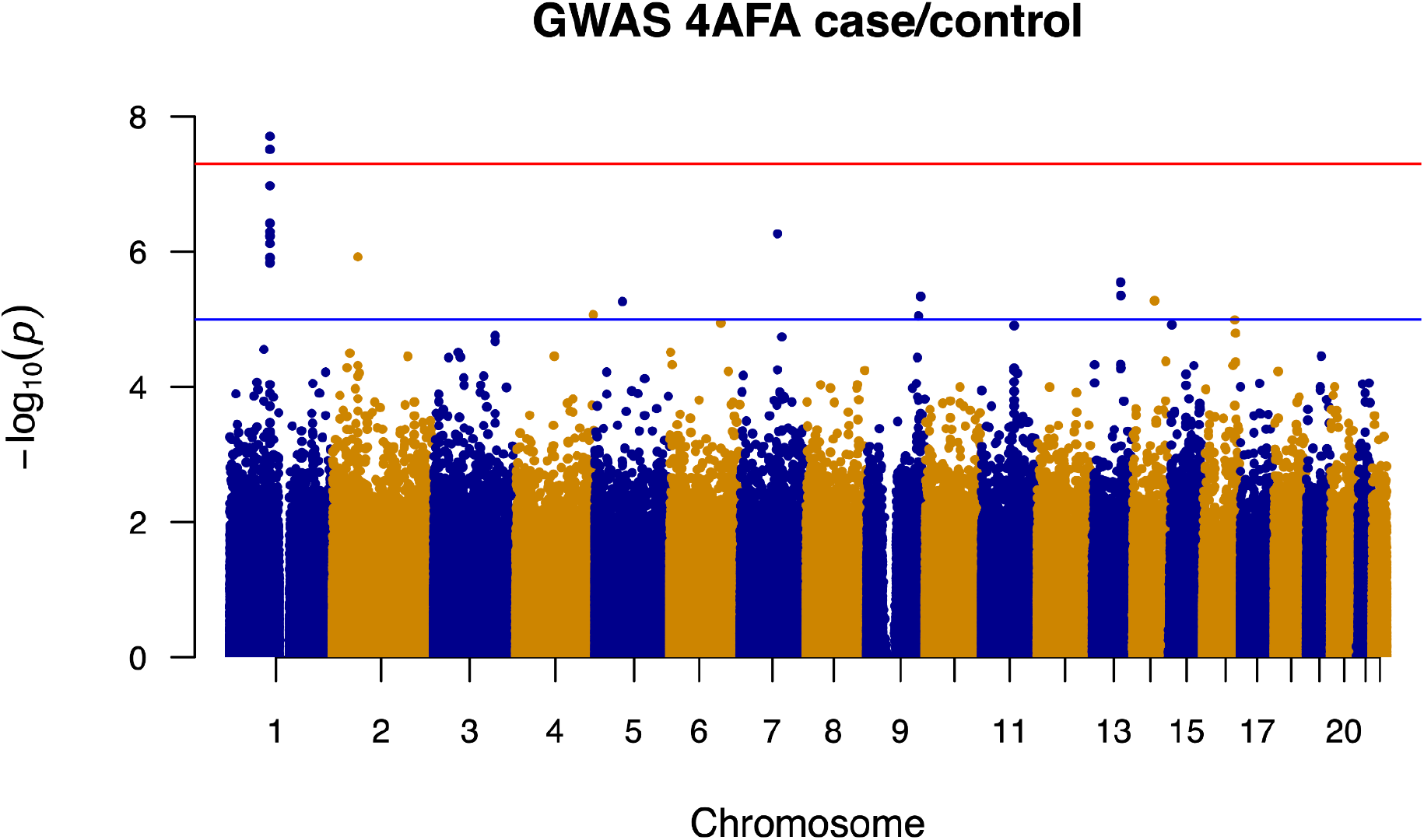
Fig. S1-S4. GWAS results for cocaine use (case) or no use (control) for (S1) 1CEU, mainly European ancestry, (S2) 4AFA, mainly African American ancestry, (S3) 5SH1, mainly Puerto Rican Hispanic ancestry, (S4) 6HS2, mainly Central and South American Hispanic ancestry. In each plot, the red line indicates statistical significance after Bonferroni correction (p < 5.5 10^-8^), blue line indicates suggestive significance (p < 1 × 10^-5^).

**Fig. S7.**
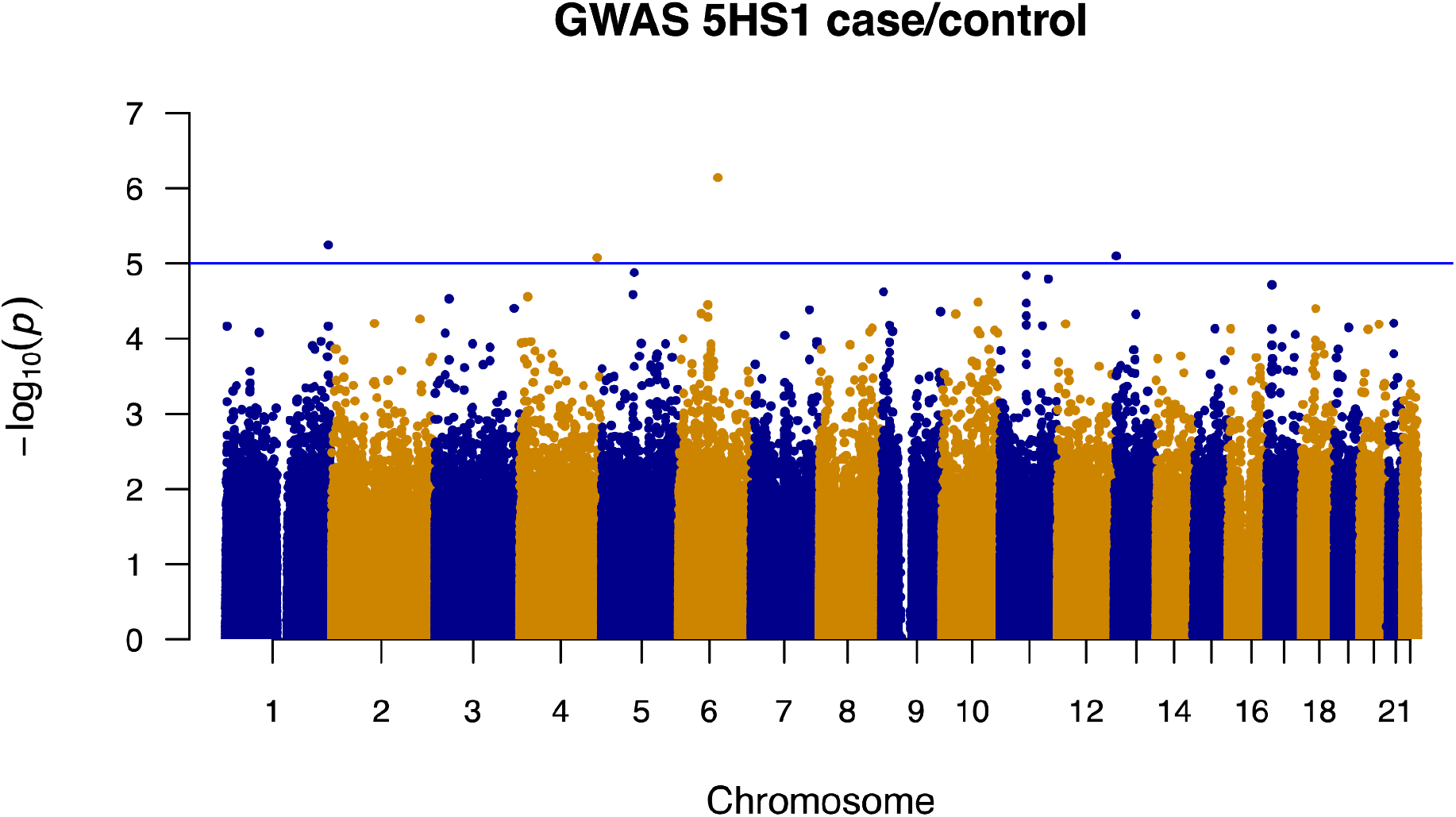
Fig. S1-S4. GWAS results for cocaine use (case) or no use (control) for (S1) 1CEU, mainly European ancestry, (S2) 4AFA, mainly African American ancestry, (S3) 5SH1, mainly Puerto Rican Hispanic ancestry, (S4) 6HS2, mainly Central and South American Hispanic ancestry. In each plot, the red line indicates statistical significance after Bonferroni correction (p < 5.5 10^-8^), blue line indicates suggestive significance (p < 1 × 10^-5^).

**Fig. S8.**
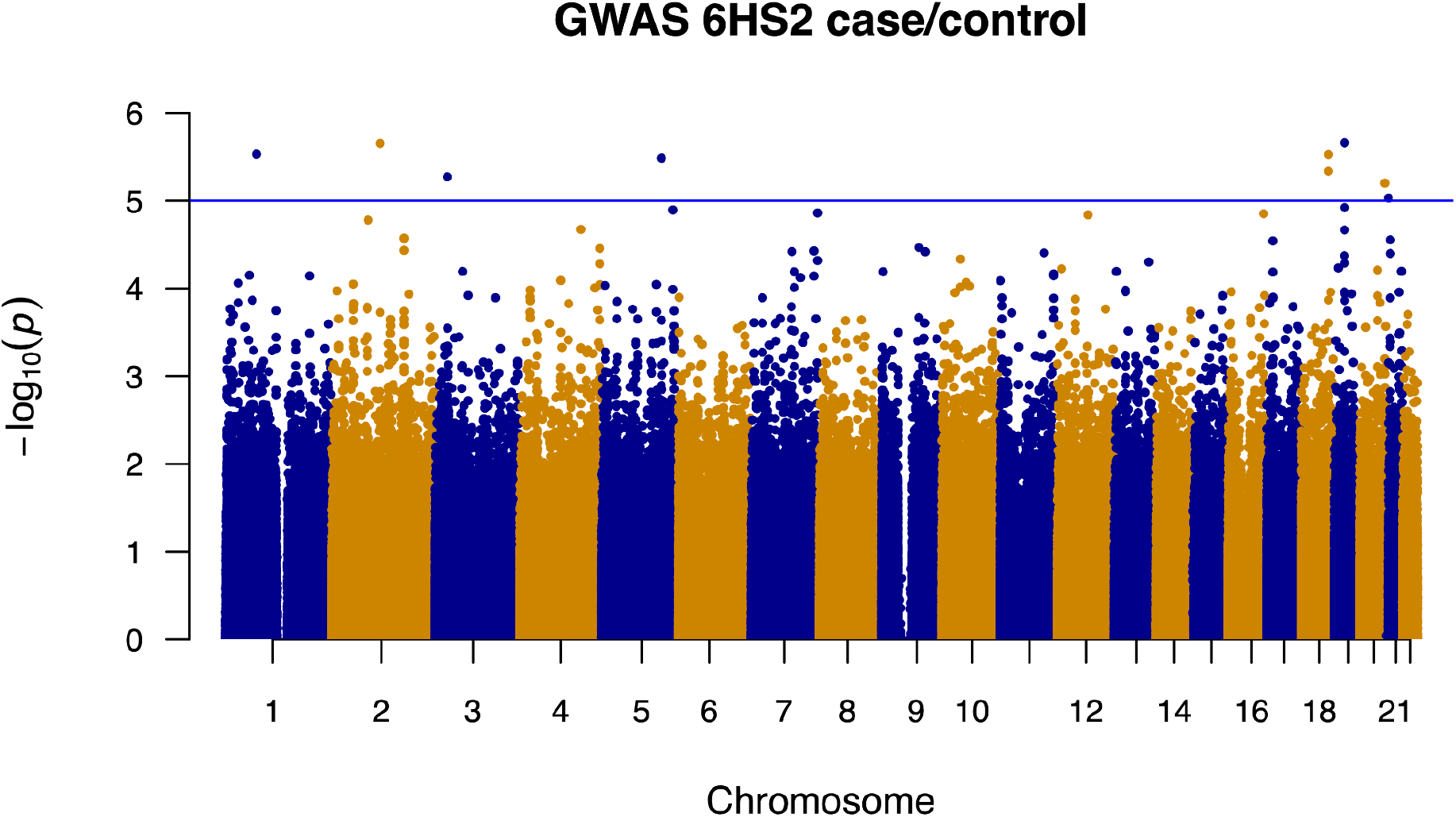
Fig. S1-S4. GWAS results for cocaine use (case) or no use (control) for (S1) 1CEU, mainly European ancestry, (S2) 4AFA, mainly African American ancestry, (S3) 5SH1, mainly Puerto Rican Hispanic ancestry, (S4) 6HS2, mainly Central and South American Hispanic ancestry. In each plot, the red line indicates statistical significance after Bonferroni correction (p < 5.5 10^-8^), blue line indicates suggestive significance (p < 1 × 10^-5^).

